# Deep Brain Stimulation for Epilepsy: Optimal Targeting and Clinical Outcomes

**DOI:** 10.1101/2025.06.23.25330152

**Authors:** Lauren A. Hart, Aaron E.L. Warren, Niels Pacheco-Barrios, Bahne H. Bahners, Savir Madan, Clemens Neudorfer, Calvin W. Howard, Melissa Chua, Young-Min Shon, Brin E. Freund, Erik H. Middlebrooks, Frederic L.W.V.J. Schaper, Sara Larivière, John D. Rolston, Andreas Horn, Garance M. Meyer

**Affiliations:** Center for Brain Circuit Therapeutics, Department of Neurology, Mass General Brigham, Harvard Medical School, Boston, MA, USA; Department of Neurosurgery, Mass General Brigham, Harvard Medical School, Boston, MA, USA; Institute of Clinical Neuroscience and Medical Psychology, Medical Faculty and University Hospital Düsseldorf, Heinrich Heine University Düsseldorf, Germany; Department of Neurology, Center for Movement Disorders and Neuromodulation, Medical Faculty and University Hospital Düsseldorf, Heinrich Heine University Düsseldorf, Germany; Department of Neurology, Samsung Medical Center, Sungkyunkwan University School of Medicine, Seoul, Korea; Department of Neurology, Mayo Clinic, Jacksonville, Florida, USA; Department of Radiology, Mayo Clinic, Jacksonville, Florida, USA; Department of Medical Imaging and Radiation Sciences, Faculty of Medicine and Health Science, University of Sherbrooke, Sherbrooke, Qc, Canada; Department of Neurology, Faculty of Medicine, University of Cologne, Cologne, Germany

**Keywords:** Deep Brain Stimulation, Epilepsy, Neuromodulation, Anterior Nucleus of the Thalamus, Centromedian Nucleus of the Thalamus

## Abstract

**Background:** Deep brain stimulation (DBS) has been investigated for patients with drug-resistant epilepsy who are not candidates for resective surgery. Because different types of epilepsy involve different brain networks, numerous DBS targets have been explored.

**Methods:** To provide a comprehensive overview of this expanding literature, we conducted a systematic review of studies for DBS in epilepsy, collecting data on surgical targets, individual disease characteristics, outcomes, and precise electrode placements. DBS electrode coordinates were gathered into a common template space and related to clinical outcomes.

**Findings:** We included 124 studies, corresponding to 1,210 patients and 20 distinct surgical targets. While the anterior (ANT) and centromedian (CM) nuclei of the thalamus remain the most studied, we also review less commonly used targets that show promise for specific forms of epilepsy and may warrant further investigation. Substantial variability in targeting strategies and electrode placement was observed within each of the target regions. Importantly, significant relationships between stimulation location and outcomes were identified for ANT– and CM-DBS. For ANT-DBS, shorter distance to the mammillothalamic tract junction was associated with greater seizure reduction on both study– and patient-level analyses (r=-0.55, p<0.001 and r=-0.51, p<0.001, respectively). For CM-DBS, localization effects may be dependent on the form of epilepsy, with stimulation of the parvocellular CM being associated with better outcomes in generalized epilepsy.

**Interpretation:** Our results emphasize the importance of accurate targeting in DBS for epilepsy. Our database and atlas of DBS targets are made publicly available, potentially serving further meta-analytical work.

**Funding:** A.H. was supported by the Schilling Foundation, the German Research Foundation (Deutsche Forschungsgemeinschaft, 424778381 – TRR 295), Deutsches Zentrum für Luft-und Raumfahrt (DynaSti grant within the EU Joint Programme Neurodegenerative Disease Research, JPND), the National Institutes of Health (R01MH130666, 1R01NS127892-01, 2R01 MH113929 & UM1NS132358) as well as the New Venture Fund (FFOR Seed Grant).

**Research in context:** *Evidence before this study:* Deep brain stimulation (DBS) is a treatment option for drug-resistant epilepsy. Given the complexity and heterogeneity of the disease, various brain networks may be candidates for targeted neuromodulation. We systematically searched the PubMed database on 07/05/2024 using the search terms “Epilep* AND (neuromodulation OR deep brain stimulation)”. Although several systematic reviews and meta-analyses evaluating DBS for epilepsy have been published in recent years, these have focused on individual surgical targets. A comprehensive synthesis of the broader and increasingly diverse landscape of DBS targeting strategies in epilepsy has been lacking. Moreover, no large-scale, systematic assessment of the relationship between electrode placement and outcomes has been conducted.

*Added value of the study:* Our systematic review provides the most comprehensive overview to date of deep brain stimulation targets for epilepsy, synthesizing data from 1,210 participants across 124 studies. We report on surgical targeting strategies, clinical rationales and outcomes for 20 distinct anatomical targets, identifying promising emerging options for specific epilepsy types. Crucially, we collected DBS electrode coordinates and pooled them across studies, revealing substantial variability in targeting strategies and electrode placement within each region. By systematically relating electrode locations to outcomes, we derive meta-analytical evidence for optimal stimulation sites within anterior and centro-median nuclei of the thalamus.

*Implications of all the available evidence:* While only the anterior nucleus is approved as a DBS target for epilepsy in the United States and Europe, our review highlights other emerging targets that warrant further investigation. Furthermore, our findings emphasize the key influence of electrode placement on outcomes and have immediate clinical relevance for DBS targeting and programming for the anterior and centro-median nuclei. Further research is needed to better define the networks modulated by stimulation (even more so for surgical targets other than the anterior and centro-median nuclei) and optimize outcomes for patients with drug-resistant epilepsy.

## Introduction

Epilepsy is among the most prevalent and debilitating neurological disorders, impacting over 50 million people worldwide^1,2^. Despite treatment with antiseizure medications, one third of patients experience persistent seizures, i.e. drug-resistant epilepsy^3^. For these individuals, surgery represents a crucial therapeutic option^4^. Resective surgery can be curative when a seizure onset zone can be identified^4^. Neuromodulation therapies such as vagus nerve stimulation (VNS), responsive neurostimulation and deep brain stimulation (DBS) may instead be considered when the seizure onset zone cannot be identified or safely removed^5^.

Among these, DBS has the longest history of research in epilepsy, with the first studies dating back 40 years^6,7^. Epilepsy is a complex and heterogeneous disease, with its various types involving distinct brain networks that may each be targeted by DBS^5,8^. Accordingly, numerous surgical targets have been explored. Of these, the anterior nucleus of the thalamus (ANT) remains the only approved DBS target for epilepsy in the US and Europe. Our systematic review aimed at providing a comprehensive overview of this expanding literature, reporting on existing surgical targets, rationales for their use, and outcomes. Of note, we collected individual patient data on disease characteristics to better elucidate the rationales for target selection.

Importantly, and as noted for other DBS indications^9,10^, the same anatomical label may refer to slightly different target locations, and stimulation to a given target may influence surrounding structures, such that it often remains unclear which exact (sub-)structures mediate the therapeutic effects of DBS. Precisely identifying the optimal stimulation sites by systematically relating stimulation locations to clinical outcomes is therefore essential for understanding DBS mechanisms and optimizing treatment. To this end, we collected coordinates of DBS electrodes for each study and – if available – also for individual patients enrolled in the studies. Notably, coordinates have been reported using various conventions over time^11^. Therefore, all coordinates were transformed into a common template space.

In addition to the findings reported in this study, we make two novel resources openly available. Namely, an atlas of DBS targets for epilepsy, with electrode coordinates for each reviewed study, and the extensive database that supported our conclusions, which consists of detailed data from 124 studies including 1,210 patients who received DBS for epilepsy across numerous international centers.

## Methods

### Data collection

We conducted a systematic review of studies of DBS for epilepsy following the PRISMA guidelines. The review was registered with PROSPERO (CRD420250649304). Since our aim was to provide an overview of targeting strategies, we included all studies in which patients received DBS for the treatment of epilepsy and in which outcomes were reported. We applied broad inclusion criteria to capture underutilized and emerging targets, which may have been thus far only described in case reports. A full description of the systematic review procedure is available in the Supplementary material.

The main variables of interest were anatomical target, epilepsy type^a^ as classified by the International League Against Epilepsy (ILAE)^11^, and outcomes, specifically the percent seizure reduction from the pre-operative baseline (SR) and/or responder status (with response being defined as SR of ≥ 50%^12^). Additionally, we collected data on lobar localization of focal seizure onset, epilepsy syndrome, epilepsy etiology, structural brain abnormalities, history of VNS, co-morbidities influencing DBS target selection, patient age, sex, seizure types, pre– and post-DBS seizure frequency, follow-up time, and study design.

Crucially, we recorded DBS target coordinates, considering both planned surgical target coordinates and active contact coordinates (i.e., the locations of the contacts delivering stimulation; see Supplementary methods). AC-PC coordinates were brought into MNI template space using the dedicated tool in Lead-DBS^13^. Information on surgical planning and intraoperative confirmation of targeting was also collected. To illustrate the different targeting approaches used in DBS for epilepsy, approximate electrode trajectories were reconstructed for example cases using the mock planning tool in Lead-DBS^14^.

### Statistical analysis

To date, there are no comparative trials of DBS targets in the same epilepsy population, such that robust comparisons of seizure outcomes across targets were not possible. Instead, we report on the effectiveness of each DBS target in the corresponding patient populations (i.e. mean SR, response rate and seizure freedom). These were calculated based on available data (missing data was not imputed; Table S3). Of note, data was analyzed both at the *study level* (when variables were reported in aggregates for the entire study sample), and at the *patient level* (for the subset of studies that reported information on individual patients). Analyses incorporating variables such as epilepsy type, which are specific to individual patients, were therefore conducted using patient-level data only.

The relationships between target or active contact coordinates and outcomes were assessed by testing for correlations with SR (Pearson’s coefficients of correlation), or by comparing responders and non-responders (two-sample t-tests). Again, this was studied both at the study level (using coordinates that surgeons used for targeting in the entire study cohort) and, when possible, at the patient level (using active contact coordinates of individual patients). P-values below α = 0.05 were considered significant.

### Data availability

The resulting database, including the clinical information and target coordinates (124 studies, N = 1,210 patients), as well as our atlas of DBS targets for epilepsy (i.e., the reconstructed electrodes compiled as a Lead-DBS dataset^14^), are available in full at https://osf.io/kpwrv/.

## Results

A total of 124 studies were included in the final analysis (see flowchart in Fig. S1 and Supplementary methods). The resulting dataset encompassed 1,210 patients (452 females, 504 males, 31.5 ± 7.5 years old), receiving DBS across 20 anatomical targets. Across studies, the average seizure reduction (SR) was 52.1 ± 51.5 % (659 responders out of 1061 with available responder status, i.e., 62.1%).

Patient-level data was available for 789 individuals (308 females, 335 males, 30.6 ± 12.1 years old), allowing for breakdown of epilepsy types. Among these, 317 had focal epilepsy, 224 multifocal epilepsy, 182 generalized epilepsy, 40 combined generalized and focal epilepsy, and 26 had an unspecified epilepsy type. The mean follow-up duration was 24.6 ± 22.3 months. Percent SR was available for 631 of these 789 patients, with a mean reduction of 54.8 ± 49.6 %, similar to study-level data. This corresponded to 532 (67.4%) responders.

Anatomical target locations are shown in Fig. 1. SR and response rate broken down per target and epilepsy type are shown in Fig. 2. Detailed results for each DBS target are provided in the Supplementary material and summarized hereafter.

**Figure 1:**
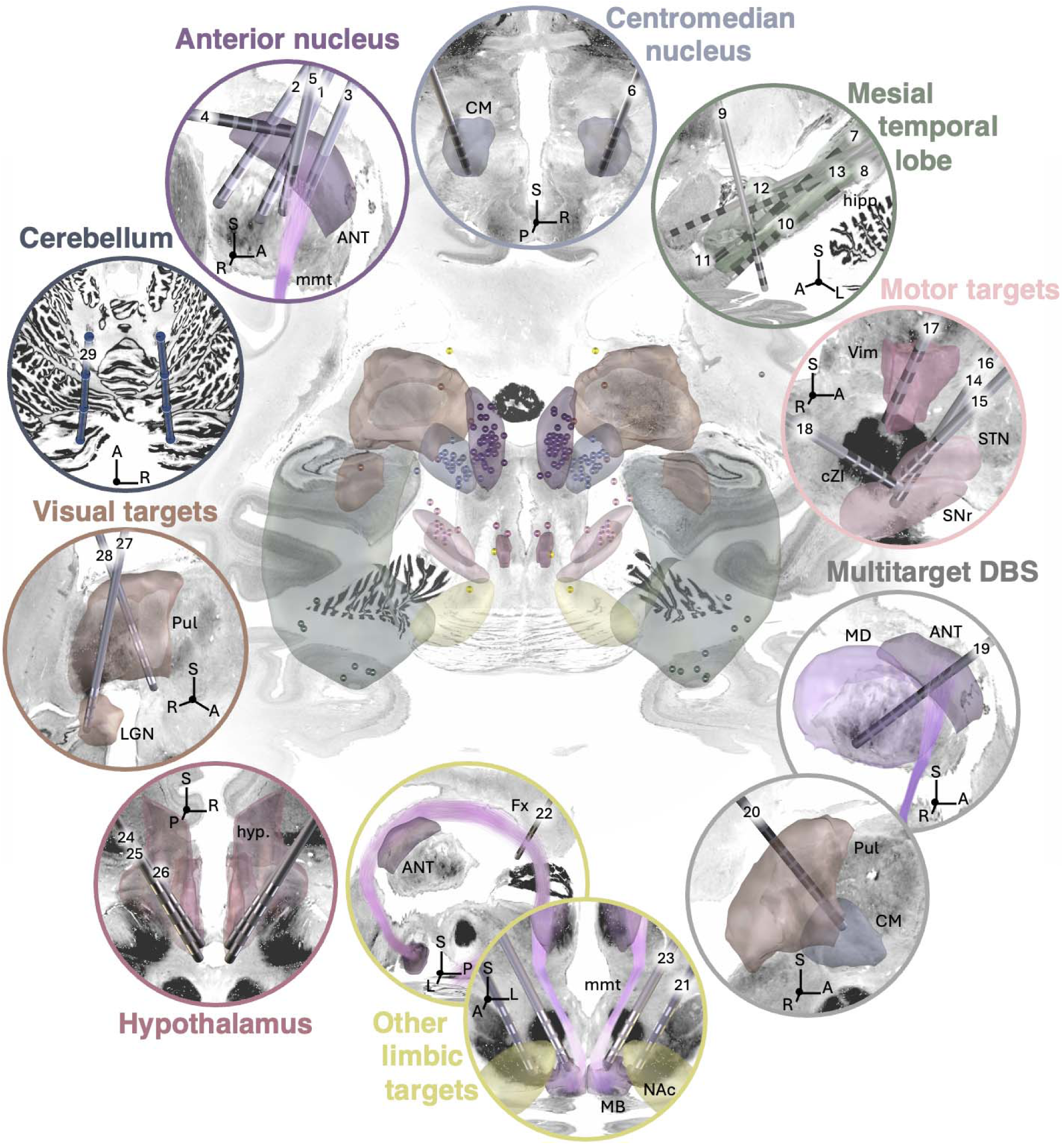
DBS targets for epilepsy. Spheres in the central panel represent study-level target coordinates. When possible, the corresponding approximate electrode trajectories were reconstructed for illustrative purposes (outer circles). The coordinates, as well as the trajectories compiled as a Lead-DBS dataset, are made openly available (https://osf.io/kpwrv/; see Table S2 for detail on the numbered trajectories). The ANT was initially targeted using an indirect transventricular approach, but more recent studies proposed extraventricular and/or direct approaches. Direct targeting uses the mmt junction (which can be visualized on FGATIR MRI sequences) as an anatomical landmark. CM implantations typically continue to rely on indirect targeting. Variability in targeting strategies is particularly pronounced for mesial temporal lobe structures. Differences in electrode placement, span, and stimulation settings likely result in large differences in the extent of stimulated structures. Different targets pertaining to the motor system have been introduced, including the cZI, Vim and STN/SNr. Several studies performed multitarget DBS, combining ANT stimulation with a secondary target. In some cases, longer electrodes with eight contacts have been used to target two adjacent thalamic nuclei within a single trajectory, such as the ANT and mediodorsal nucleus, or CM and pulvinar. Other limbic targets included the NAc, mmt and fornicodorsocommissural tract, located near the crus of the fornix. Studies of hypothalamic DBS targeted the posterior part of the structure. Targets pertaining to the visual system included the pulvinar and LGN. Finally, the cerebellum has been targeted with four-button electrodes placed at its superomedial surface. ANT: anterior nucleus of the thalamus; CM: centro-median nucleus of the thalamus; cZI: caudal zona incerta; Fx: fornix; Hipp.: hippocampus; hyp.: hypothalamus; LGN: lateral geniculate nucleus; MB: mammillary bodies; MD: mediodorsal nucleus of the thalamus; mmt: mammillothalamic tract; NAc: nucleus accumbens; Pul: pulvinar; STN: subthalamic nucleus; SNr: substantia nigra pars reticulata; Vim: ventral intermediate nucleus of the thalamus.

**Figure 2:**
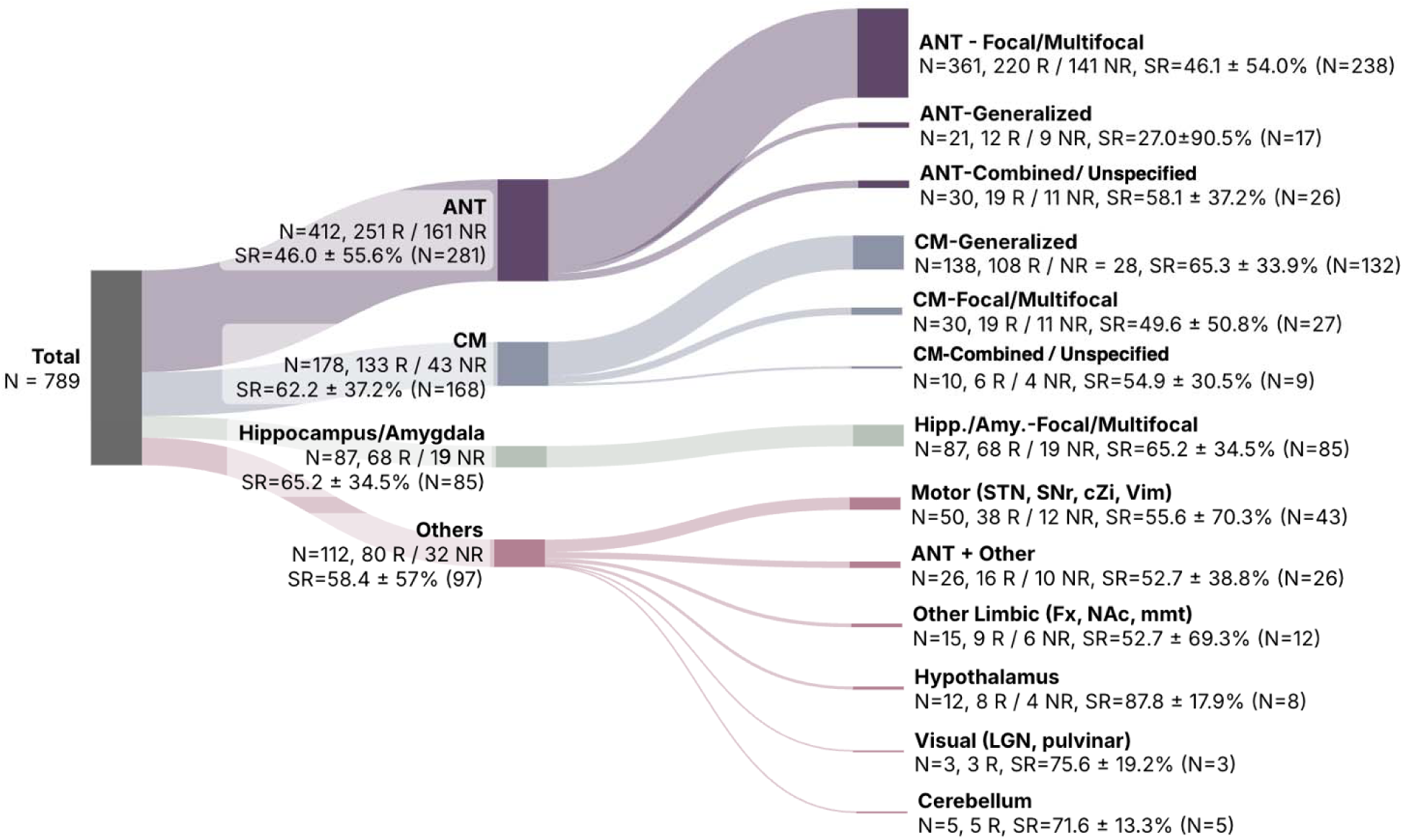
Number of patients and outcomes broken down by DBS target and epilepsy type (analysis based on patient-level data). Numbers given in parentheses represent how many patients contributed to the calculation of average seizure reduction for each subgroup, reflecting the missing data points that were excluded from the analysis (e.g., for ANT, responder status was available for 412 patients, but average seizure reduction was calculated only across 281 patients with corresponding data). NR: non-responders; R: responders; SR: seizure reduction.

### Anterior nucleus of the thalamus

Sixty studies (48.4%) reported on ANT-DBS, including a total of 801 patients (66.2% of all patients analyzed here). The mean SR was 46.3 ± 19.1%, with 375 patients classified as responders out of 684 with available responder status (54.8%; average FU time: 22.4 ± 16.9). Two randomized-controlled trials (RCTs) were conducted: the SANTE trial^15^ (N = 110) reported a median SR of 40.4% (vs. 14.5% in controls) at the end of the 3-month blinded phase, while a smaller RCT by Herrman et al.^16^ (N = 18) found a 22% SR at the end of the 6-month blinded phase, but no significant benefit over controls. Patient-level data showed that most patients had focal or multifocal epilepsy, and that responder rates were generally consistent across epilepsy types, although SR was numerically lower for patients with generalized epilepsy (Fig. 2 and Supplementary results). Electrodes were implanted using a frontal transventricular, extraventricular, or posterior parietal extraventricular approach (Fig. 1). Leveraging coordinates in our database, we tested two a priori hypotheses from the published literature. First, we could confirm that more anterior target coordinates were associated with better outcomes (study level: 43 studies, r = 0.34, p = 0.044), as proposed by Lehtimäki et al.^17^. Second, we found that outcomes were most favorable when stimulation was delivered near the junction of the ANT and mammillothalamic tract, as proposed by Schaper et al.^18^ (study level target coordinates: 43 studies, r = –0.55, p < 0.001; patient level active contact coordinates: 5 studies, 52 patients, r = – 0.51, p < 0.001; Fig. 3 and Supplementary results).

**Figure 3:**
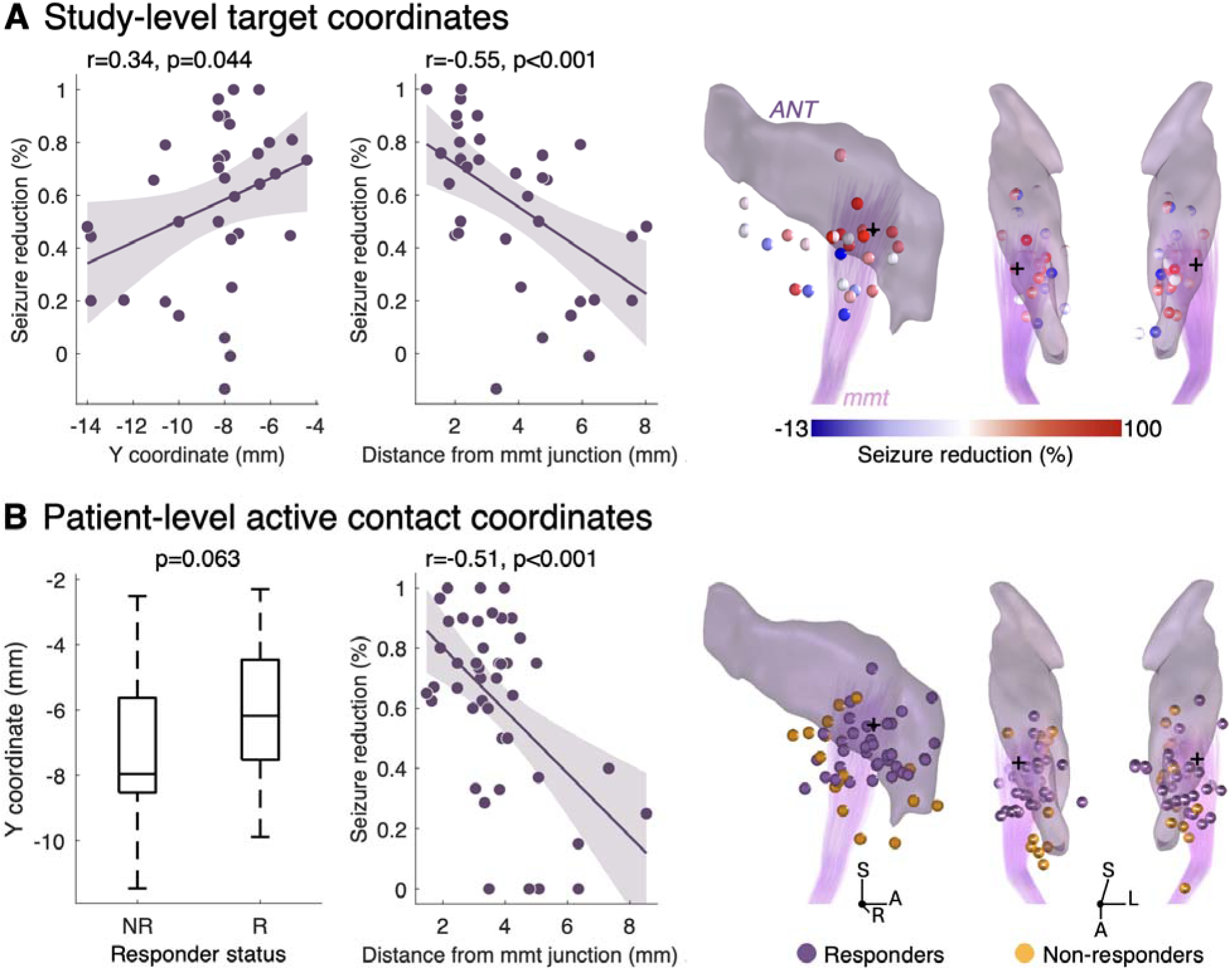
Relationship between electrode position and outcomes for ANT-DBS. (A) At the study level (43 studies), more anterior target coordinates (i.e., the intended, functional coordinates used for surgical targeting in all patients of a given study) and shorter Euclidean distance from the mammillothalamic tract junction (mmt; as identified in MNI space and represented with black crosses) were associated with better seizure outcomes. Of note, significantly more anterior electrode placement was found for studies performing direct targeting using the mmt as an anatomical landmark as compared to earlier studies relying on indirect targeting (see Supplementary results for detail). (B) Five studies reported patient-specific active contact coordinates (i.e., coordinates for the specific contacts delivering stimulation in each patient; 52 patients)^19–23^. When pooling data across these studies, there was a trend for more anterior active contacts in responders compared to non-responders. Similar to what was observed at the study level, shorter distance from the mmt junction was associated with better outcomes.

### Centromedian nucleus of the thalamus

Thirty-two studies (25.8%) performed CM-DBS, including 208 patients (17.2% of all patients analyzed here). The mean SR was 64.2 ± 20.0%, with 76.4% of patients classified as responders (136 out of 178 with available responder status; average FU time: 20.7 ± 27.0). Two RCTs reported median SRs of 30%^24^ (not significantly different from the 8% SR off-stimulation) and 35%^25^ (with higher responder rate in the stimulation vs. control group based on electrographic seizures). Patient-level data showed that most patients had generalized epilepsy (Fig. 2), most often in the form of Lennox-Gastaut or –as reported in the corresponding publications– “Lennox-Gastaut like” syndrome.

Electrode implantation typically relied on indirect targeting, sometimes complemented by MER or intraoperative stimulation and EEG recording of recruiting responses (Fig. 1). Overall, available data suggests that greater SR is associated with stimulation of the anterior and inferolateral (parvocellular) part of the CM in patients with generalized epilepsy. Conversely, for multifocal epilepsy, greater SR may be associated with stimulation of the mediodorsal nucleus, although this result should be interpreted with caution given the small sample size (Fig. 4 and Supplementary results).

**Figure 4:**
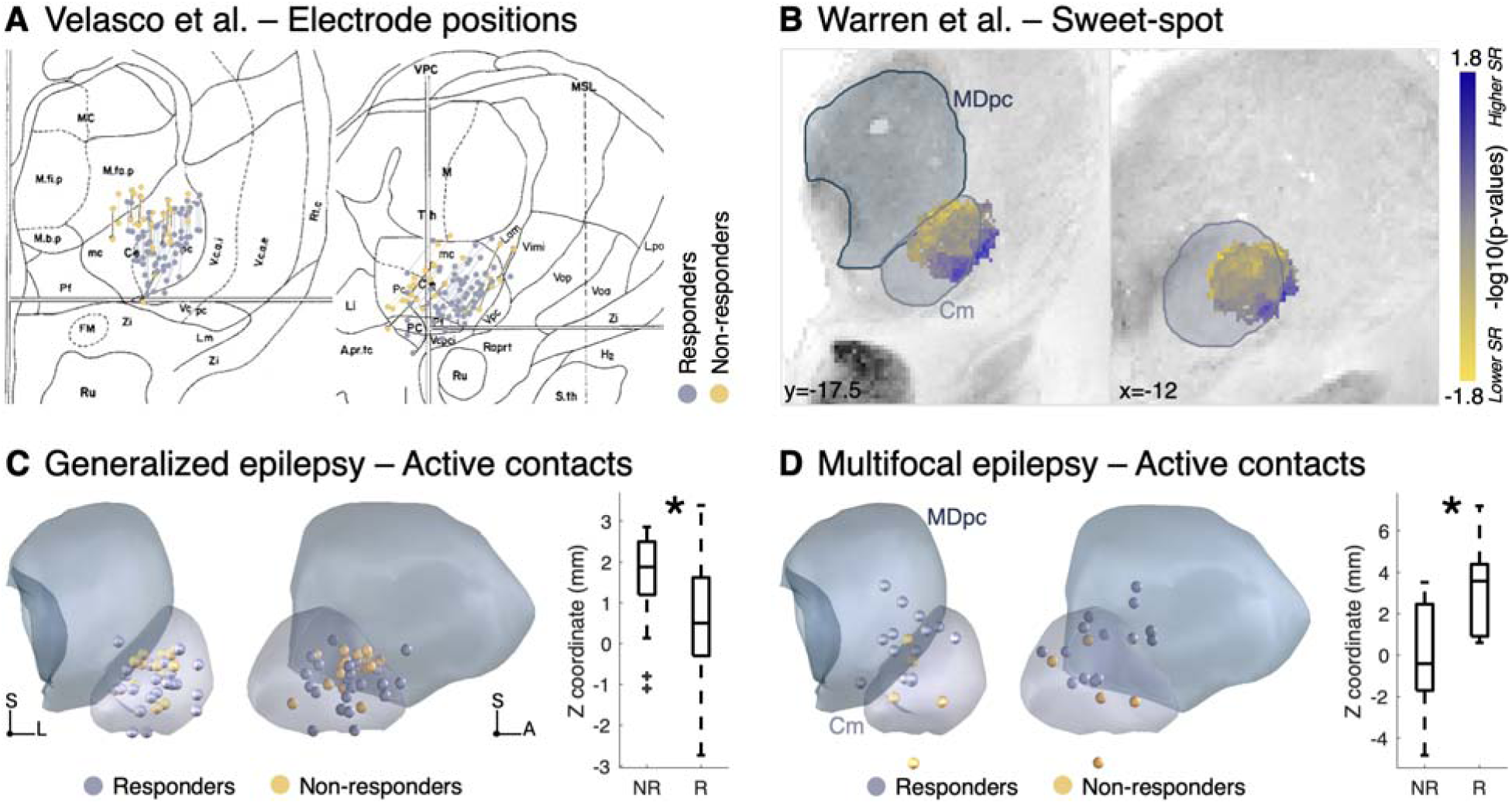
Relationship between electrode position and outcomes for CM-DBS. Since CM-DBS mostly relied on indirect targeting, there was limited variance in targeting coordinates at the study level (26 studies), such that no significant correlations with SR were found. Analysis of active contact positions in patients with generalized epilepsy suggested that greater SR is associated with stimulation of the anterior and inferolateral (parvocellular) part of CM. Panels A and B represent findings from previous published studies, while panels C and D reflect results from the present analysis. Panel A represents electrode positions for responders and non-responders as reported by Velasco et al. (coronal and sagittal views adapted from ^26,27^, total N = 26; precise coordinates could not be extracted for these studies). Panel B shows the original sweet-spot map derived by Warren et al.^28^ in patients with Lennox-Gastaut syndrome (coronal and sagittal views, N = 19). Positive values represent voxels whose stimulation was associated with better outcomes, (blue), while negative values represent voxels whose stimulation was associated with less favorable outcomes (yellow). (C) Patient-level active contact coordinates were only available for two studies in our database (Son et al. and Warren et al.^28,29^, total N = 33). Combining data from these studies, active contacts were situated significantly more ventral in responders than non-responders when considering patients with generalized epilepsy (t(40) = – 2.5, p = 0.018, with no significant difference on the other axes; anterior and lateral views). (D) Interestingly, and although this should be interpreted with caution, this pattern was reversed in the patients with multifocal epilepsy reported by Son et al.^29^, with responders’ active contacts being significantly higher (t(17) = 2.8, p = 0.014, with no significant difference on the other axes) and mainly located in the adjacent mediodorsal nucleus of the thalamus. Note that all coordinates were plotted to the same hemisphere in this figure, consistent with the original publications.

### Other targets

Beyond ANT and CM-DBS, a variety of other targets have been explored for epilepsy, including the hippocampus and mesial temporal lobe structures, motor (STN, SNr, cZI, Vim) and limbic structures (NAc, mmt, fornicodorsocommissural tract), hypothalamus, visual thalamic nuclei (LGN, pulvinar), and cerebellum (Fig. 1 and 2). Among these, hippocampal DBS was the most studied (15 studies, N = 89 patients), with a responder rate of 78.2% (68 of 87 with available data) and a mean SR of 64.5 ± 19.1%. Motor targets—particularly the STN/SNr and cZI—showed promising results in patients with motor seizures or myoclonic epilepsies. Multitarget DBS strategies (e.g., ANT+CM) yielded moderate response rates, while stimulation of other limbic structures, hypothalamus, and cerebellum also showed favorable outcomes in small cohorts. Studies varied widely in targeting strategies and patient populations, and the influence of precise stimulation location on outcomes remains poorly characterized for these targets.

In line with the journal’s formatting restrictions, we provide an extensive summary about these targets, as well as additional views of electrode placements, as Supplementary results and Fig. S3-7.

## Discussion

DBS has been investigated as a treatment option for patients with drug-resistant epilepsy who are not candidates for resective surgery. In this work, we summarize the DBS targets investigated to date, the clinical rationales for their use, and the associated outcomes. While ANT and CM remain the most studied, we highlight less commonly used targets –such as motor regions– that show promise for specific forms of epilepsy and may warrant further investigation. Crucially, our analysis reveals substantial variability in targeting strategies and electrode placement within each of the target regions. By meta-analytically pooling data across studies, two key insights could be derived regarding the influence of these factors on outcomes. First, for ANT-DBS, we show that stimulation closer to the ANT-mmt junction is associated with greater seizure reduction, now providing meta-analytic, larger-scale evidence in support of previous hypotheses^18^. Second, for CM-DBS in generalized epilepsy, stimulation of the parvocellular part of the nucleus was linked to better outcomes. Locations of optimal stimulation sites remain largely unknown for the other surgical targets. Our database (N = 124 studies; 1,210 patients) and atlas of DBS targets for epilepsy are made publicly available for further research.

### DBS targets for epilepsy

Our systematic review showcases the variety of DBS targets that have been investigated. Randomized controlled trials have only been performed for ANT^15,16^, CM^24,25^, hippocampus^30^ and NAc^31^; other targets were evaluated in small and/or open label studies. Only ANT-DBS is currently approved for the treatment of epilepsy in the United States, Europe and Canada (Fig. 5).

**Figure 5:**
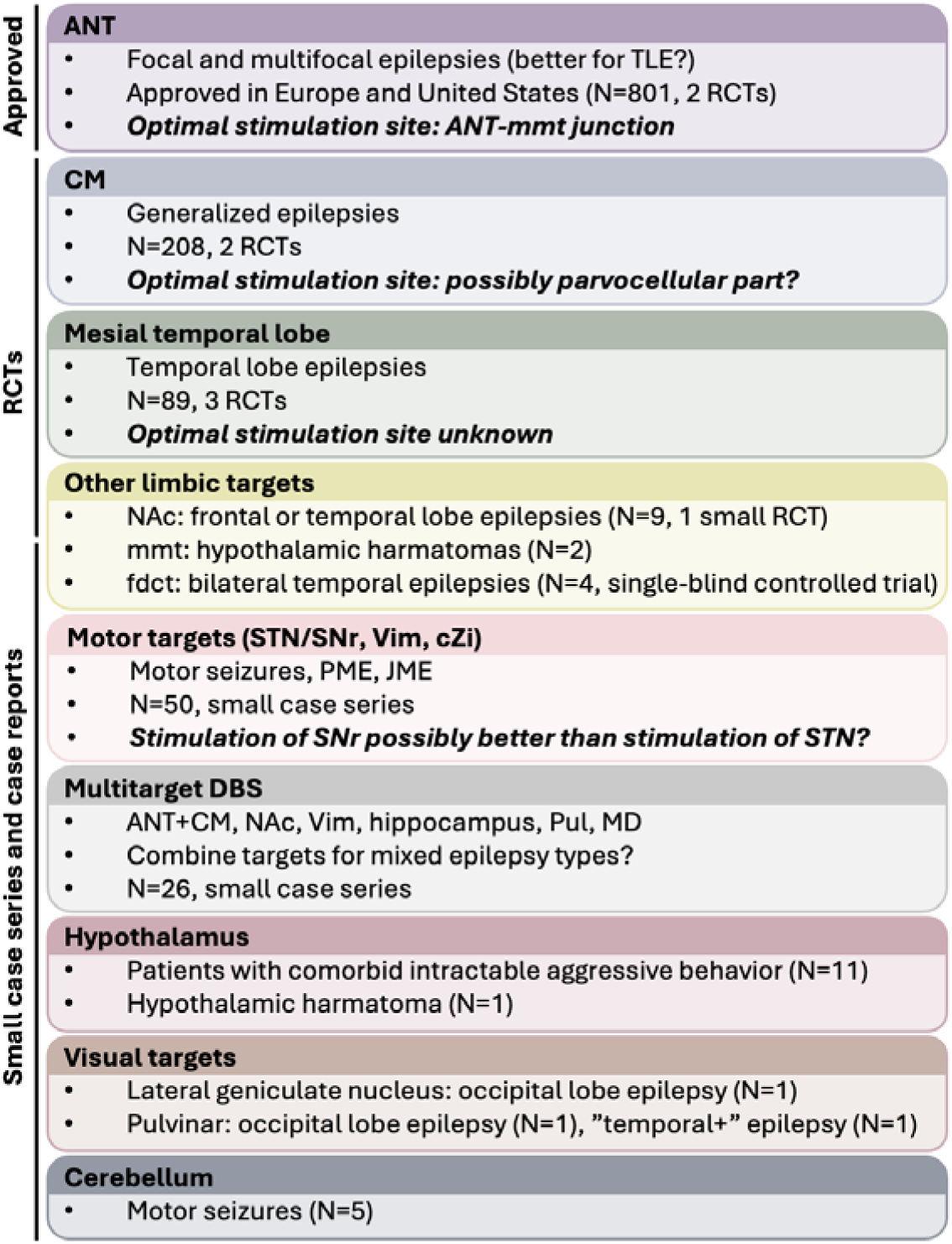
Summary of existing surgical targets, indications, optimal stimulation sites and level of evidence. JME: juvenile myoclonic epilepsy, PME: progressive myoclonic epilepsy, TLE: temporal lobe epilepsy,

Typically, neuromodulation is thought to reduce seizures by modulating the brain regions or networks that may contribute to their generation or propagation^8^. Hence, novel targets have been selected based on their position in such networks, and sometimes based on animal studies demonstrating potential efficacy in reducing seizures^5^. The ANT is a key node of the Papez circuit^32^, which has been implicated in the propagation of seizures^33^ and interconnects with mesial temporal and cingulate regions. High frequency stimulation of ANT has been shown to produce widespread activation within the limbic and default mode networks^34^. Similarly, the NAc has strong connections to frontal and mesial temporal regions and has been investigated for focal and multifocal epilepsies involving these regions^35^. The mesial temporal lobe, and particularly the hippocampus, is highly prone to seizures. It was therefore hypothesized that high-frequency stimulation of the hippocampus might suppress seizure activity by mimicking the lesional effect of an ablation or resection^36,37^. Interestingly, recent data suggests efficacy of hippocampal DBS for epilepsies beyond that of the temporal lobe^38^. Stimulation of the fornico-dorsocommissural tract relies on a similar rationale^39,40^, although low-frequency stimulation was applied in this case.

The CM is part of the thalamic activating system^41,42^, which influences large parts of the cortex. This key position, as well as early animal work^41^, has motivated its use for generalized epilepsy^43^. However, some studies have targeted the CM for focal epilepsies originating from frontal regions.

The hypothalamus has been used primarily in patients suffering from epilepsy with comorbid intractable aggressive behavior^44–47^. The pulvinar and LGN have been used for occipital epilepsies, consistent with their involvement in the visual system and structural connections with the occipital lobe. The widespread connections of the pulvinar also motivated its use for “temporal+” epilepsy, where seizures originate in the temporal lobe but propagate to adjacent regions^48–51^.

The STN, STN/SNr and Vim have been used for epilepsies originating from central regions or involving seizures with motor semiology, as well as for progressive and juvenile myoclonic epilepsies (see for review ^52^). Still, while all these structures are part of motor networks, they differ in their connectivity patterns, and there is currently insufficient data to assess which one is most promising. Similarly, Velasco et al.^53^ targeted the superomedial surface of the cerebellum for intractable motor seizures. The cerebellum participates in whole-brain motor networks^54^ and has been targeted mainly based on animal studies demonstrating inhibition of seizures^6,55,56^. It should be noted that cerebellar DBS was investigated in the 1970s and 1980s with conflicting results^57^. Finally, a few other historical targets have not yet been reinvestigated in the “modern” era of DBS, such as the caudate^58^ or locus coeruleus^59^.

Interestingly, institutions have started offering off-label personalized target selection based on the patient’s epilepsy type and corresponding hypotheses about involved networks^50,51,60^. Overall, while a small number of studies directly investigated the networks implicated in DBS response^28,44,61^, the mechanisms of DBS remain incompletely understood. Neuromodulation may also improve seizures by mechanisms that are independent of seizure type^62^, for example by modulating regions that regulate overall cortical arousal^63^.

### Factors influencing outcomes

Our results emphasize that while DBS can be effective, not all patients benefit, and seizure freedom is rarely achieved. Patient characteristics and disease variables likely contribute to outcome variability^28^, but were inconsistently reported (Table S3). Given the high anatomical complexity of most DBS target regions (e.g., ^42,64,65^), electrode placement likely constitutes another important source of variance in outcomes^66,67^, as it is the case for other DBS indications^68^. Indeed, there is variability in surgical targeting coordinates and approaches for the same target region (Fig. 1), and final electrode placement in each patient may differ substantially from the average planned surgical target (Fig. 3 and 4; ^15,26–29,69^). Postoperative selection of active contacts further increases the variability introduced by differences in electrode placement.

The meta-analytical evidence gathered in the present study revealed an association between shorter distance from the ANT-mmt junction and greater seizure reduction in patients receiving ANT-DBS, both when analyzing study-level targeting coordinates and when analyzing patient-level active contact coordinates (R^2^ = 0.26 and 0.30; Fig. 3). This is consistent with results from previous individual studies (^18,20,70,71^; Table S1), and may emphasize the importance of white matter involvement for DBS effects^72,73^. These findings also highlight the relevance of direct targeting approaches that rely on individualized identification of the mmt as an anatomical landmark on specialized MRI sequences^74^. Limited evidence is available for CM-DBS, but current data suggests that stimulation of the parvocellular (anterior and inferolateral) part of the nucleus^42,75^, which has stronger projections to the premotor and precentral cortices^76^, may be associated with greater seizure reduction^26–29,77^ in patients with generalized epilepsy. In line with this result, greater seizure reduction has been associated with connectivity from stimulation sites to a reticular system network encompassing sensorimotor and supplementary motor cortices as well as the brainstem and cerebellum^44^. An important caveat is that most of the currently available data comes from patients with Lennox-Gastaut syndrome, such that it remains unclear whether these conclusions would apply to idiopathic generalized epilepsy. Indeed, the data by Son et al. suggests that optimal stimulation sites within the same target region may vary according to epilepsy type, as patients with multifocal epilepsy who responded to CM-DBS were shown to have active contacts placed dorsally in the adjacent MD thalamus^29^.

Location of optimal stimulation sites remain largely unknown for target structures other than ANT and CM. We found considerable variability in targeting strategies for mesial temporal lobe structures (Fig. 1). Differences in placement, electrode span, and stimulation settings likely result in large differences in the extent of stimulated structures, and there is insufficient data to assess the influence of these factors to date. This issue is further complicated by the presence of hippocampal sclerosis in many patients, which has been linked to worse outcomes of hippocampal DBS^79,80^. Similarly, for the STN/SNr target, it remains uncertain whether the STN or SNr represents the optimal stimulation site^52^, although small studies reported that stimulation of SNr contacts was more efficient than stimulation of STN contacts^81,82^.

Of note, there is currently very little data available to compare the efficacy of different targets within the same population of patients. Only the study by Kowski et al. performed blinded head-to-head comparison of NAc vs. combined ANT and NAc-DBS in four patients^83^. Finally, stimulation parameters other than choice of active contact, such as frequency and cycle duration, may also influence outcomes^84^.

### Limitations

Our systematic review has several limitations. To provide a complete overview of current DBS targets for epilepsy, we used broad inclusion criteria. This means that most of the included data is from small, uncontrolled studies with heterogeneous patient populations, sample sizes, study designs, and follow up times (with only few studies providing long-term data^8885^). Hence, outcomes could not be robustly compared across targets (i.e., by conducting a meta-analysis). Other limitations stem from weaknesses in reporting. For example, epilepsy type was not always reported and did not always adhere to ILAE guidelines, even in the most recent studies, highlighting the need for more standardized reporting. Information on epilepsy syndromes, etiology, and history of VNS was often incomplete, largely preventing us from assessing the influence of these variables. For some target groups, SR was not reported especially for non-responders, precluding robust calculation of average within these subgroups. Finally, we must emphasize that SR is a noisy measure (typically based on seizure diaries)^25^, which might be more or less reliable in specific patient populations and may have been calculated in different ways across studies (e.g., based on all seizure types or only most disabling seizures). When it comes to coordinates, the available information was vastly heterogeneous (e.g., coordinates given in AC-PC or MNI space, or corresponding to the tip or middle point on the electrode). However, we believe that identifying relationships between stimulation locations and outcomes despite this noise in the data only underscores the strength of these associations. Another limitation is that our systematic review did not include data on responsive neurostimulation or chronic cortical subthreshold stimulation, which apply different stimulation modes and were not suitable for direct comparison with DBS, falling outside the scope of the present work. However, studies of subcortical (thalamic) responsive neurostimulation are gaining traction^86^, further blurring the lines between these two neuromodulation techniques.

### Conclusions

In conclusion, our systematic review maps the current landscape of surgical targets in DBS for epilepsy. By meta-analytically pooling data across studies, we could identify key insights regarding optimal stimulation location in both ANT– and CM-DBS, with immediate clinical relevance for DBS targeting and programming. Moreover, we highlight the promise of rarely used targets such as (sub)thalamic motor regions.

## Acknowledgements

A.H. was supported by the Schilling Foundation, the German Research Foundation (Deutsche Forschungsgemeinschaft, 424778381 – TRR 295), Deutsches Zentrum für Luft-und Raumfahrt (DynaSti grant within the EU Joint Programme Neurodegenerative Disease Research, JPND), the National Institutes of Health (R01MH130666, 1R01NS127892-01, 2R01 MH113929 & UM1NS132358) as well as the New Venture Fund (FFOR Seed Grant). A.E.L.W. was supported by funding from the King Trust Postdoctoral Research Fellowship Program (Bank of America Private Bank, Trustee), the Program for Interdisciplinary Neuroscience at Mass General Brigham, the LGS Foundation, the Pediatric Epilepsy Research Foundation (PERF), and Spaulding Rehabilitation’s Innovative Treatments in Disorders of Consciousness grant program. F.L.W.V.J.S. was supported by the National Institutes of Health and American Epilepsy Society. BHB gratefully acknowledges support by the Prof. Dr. Klaus Thiemann Foundation (Parkinson Fellowship 2022). These funding sources had no role in study design, data collection, data analysis, data interpretation, or writing of the report.

## Author contributions

G.M.M. and A.H. conceived and designed the study. G.M.M and L.A.H. performed the systematic review, collected and verified the data, and conducted the analyses. G.M.M and L.A.H. independently accessed and verified the underlying data. A.E.L.W., N.P., B.H.H., S.M. assisted with data extraction and interpretation. C.N., C.W.H., M.C., Y.-M.S.., B.E.B., E.H.M., F.L.W.V.J.S., S.L. and J.R. contributed to study design, data interpretation, and manuscript revision. L.A.H drafted the manuscript with input from G.M.M. and A.H. All authors had full access to the data, contributed to data interpretation, reviewed the manuscript, and approved the final version for submission. All authors had final responsibility for the decision to submit for publication.

## Footnotes

a Because many of the included studies were published before publication of the ILAE guidelines in 2017, classification was sometimes inconsistent, and most studies made a distinction between focal and multifocal epilepsy. Hence, this distinction was kept in our analysis, leading to classification of epilepsy type as focal, multifocal, generalized, combined or unknown.

