## Supplementary material for "Deep Brain Stimulation for Epilepsy: Optimal Targeting and Clinical Outcomes"

Hart et al.

**Supplementary material - Deep Brain Stimulation for Epilepsy:**

**Optimal Targeting and Clinical Outcomes**

**Contents**

Supplementary Methods	p. 2

Figure S1: PRISMA flow chart	p. 5

Supplementary Results	p. 6

Figure S2: Seizure reductions across targets	p. 14

Figure S3: Electrode positions for DBS studies targeting the	p. 15
mesial temporal lobe

Figure S4: Electrode positions and targeting	coordinates for	p. 16
DBS studies investigating motor targets

Figure S5: Electrode positions and targeting	coordinates for	p. 17
DBS studies investigating other limbic targets

Figure S6: Electrode positions and targeting coordinates for	p. 17
DBS studies targeting the hypothalamus

Figure S7: Electrode positions and targeting	coordinates for	p. 18
DBS studies investigating visual targets

Table S1: Summary of studies investigating the relationship	p. 19
between electrode placement and treatment outcomes

Table S2: List of electrode trajectories	p. 21

Table S3: Summary of available data	p. 23

List of included studies	p. 24

References for supplementary material	p. 33

**Supplementary methods**

We conducted a search of the PubMed database on 07/05/2024 using the following search query: “Epilep*[Title/Abstract] AND (neuromodulation[Title/Abstract] OR deep brain stimulation [Title/Abstract])”. We included case reports, case series, pilot investigations, open-label studies, retrospective and prospective studies, registry-based analyses, and randomized controlled trials published in English since 1976. The systematic review was registered on PROSPERO (CRD420250649304) and followed the PRISMA guidelines.

*Study selection*

Studies were eligible for inclusion if they utilized deep brain stimulation (DBS) as a therapeutic intervention for epilepsy and reported seizure outcomes (i.e., seizure reduction and/or responder status). Studies were excluded if DBS was used to treat a comorbidity other than epilepsy (for example, case reports of GPi-DBS used to treat dystonia with comorbid epilepsy, with no expected benefit on seizures) or if the primary therapeutic interventions were limited to responsive neurostimulation (RNS), vagus nerve stimulation (VNS), or chronic cortical subthreshold stimulation without concurrent DBS. While RNS and chronic cortical subthreshold stimulation represent invasive brain stimulation methods like DBS, their mechanistic effects differ from DBS. In the case of RNS, stimulation is applied periodically in response to sensing specific electrophysiological activities^1^, while DBS is typically applied chronically without reference to ongoing brain activity. As name indicates, chronic cortical subthreshold stimulation uses lower voltages or currents that are not believed to result in the activation of neural elements, and is typically applied to the cortical seizure onset zone^2^, while DBS is typically applied at subcortical targets that are believed to be important nodes of the broader epileptic network^3^. Of note, we only included studies in which chronic stimulation was delivered using a fully implantable system (IPG), meaning that we de facto excluded older DBS studies performed with externalized systems, whose outcomes may have been harder to compare with modern literature in any case. Additional exclusion criteria included conference abstracts, preprints, commentaries, and animal studies. Cohorts with multiple publications were included only once, keeping the publication with the most complete information (see below).

*Clinical data extraction*

The following variables were extracted from each study when available: DBS target (as described by authors), epilepsy type, lobar localization of focal seizure onset, epilepsy syndrome, epilepsy etiology, structural abnormalities or lesions, history of VNS, co-morbidities influencing DBS target selection, patient age at the time of DBS, sex, seizure types, pre- and post-DBS seizure frequency, follow-up time, percent seizure reduction from the pre-operative baseline (SR), responder status (with patients being considered as responders if they achieved a SR of ≥ 50%^4^), and study design (randomized controlled trial or other). When possible, this information was collected for each individual patient, rather than at the study level (see flowchart; Fig S1). Additionally, total number of patients, mean and standard deviation of SR were extracted or calculated for each study.

Epilepsy types were classified using the guidelines from the International League Against Epilepsy (ILAE^5^). However, because many of the included studies were published before publication of these guidelines in 2017, classification was sometimes inconsistent, and most studies made a distinction between focal and multifocal epilepsy. Hence, this distinction was kept in our analysis, leading to classification of epilepsy type as focal, multifocal, generalized, combined or unknown. If the epilepsy type was not specified in a paper, it was reported as unspecified. Classification of seizures was found to be very heterogeneous across studies. Nevertheless, when available, we collected data on seizure types as stated in the original studies. To align with current ILAE standards, terminology has been harmonized where feasible. For example, “partial” has been standardized to “focal,” “simple partial” to “focal aware”, and “complex partial” to “focal impaired awareness”^5^. Additionally, when available, epilepsy etiologies were classified in accordance with the ILAE guidelines, categorized as genetic, structural, infectious, immune, metabolic, idiopathic or unknown^5^. Despite the absence of a formal ILAE classification^6^, information on epilepsy syndrome was also collected. This information is reported as given in the initial studies and may reflect evolving diagnostic criteria and classification schemes (e.g., some patients were described as having “Lennox-Gastaut-like” syndrome, likely because such patients would no longer meet current criteria for Lennox-Gastaut syndrome but were previously classified as such). In cases where syndromes were not explicitly reported in the studies, they were inferred -when possible- based on characteristics such as seizure types, EEG findings, and imaging features. In such cases, classification was validated based on consensus between two raters (L.A.H., A.W.).

*Targeting coordinates*

DBS target coordinates were extracted both at the study level and -when available- at the patient level. The following coordinates were considered, with the following order of preference:

- Active contact coordinates (that is, the electrode contact(s) actively delivering stimulation), in MNI or AC-PC space
- Target coordinates (that is, the anatomical location selected for electrode placement), in MNI or AC-PC space. Of note, target coordinates may refer to the location targeted for the tip, most distal contact, or middle of the DBS electrode; this information was collected when available.
- Coordinates of most distal contact, as estimated based on images shown in the publication. Publications were required to show at least two planes (axial, coronal and/or sagittal) in order to estimate coordinates with sufficient precision.

AC-PC coordinates were converted to MNI space using the dedicated tool in Lead-DBS, which accounted for the different conventions in reporting (e.g., coordinates reported with the anterior commissure, mid-commissural point or posterior commissure as a reference)^7^. Additionally, the following information on targeting was collected: use of direct or indirect targeting, use of additional information to help surgical planning or intraoperative confirmation of targeting (such as micro-electrode recordings, intraoperative EEG or stimulation), and for ANT, use of a transventricular or extraventricular approach. Finaly, active contact was recorded when this information was available.

To better showcase targeting approaches, approximate electrode positions were reconstructed using the mock planning tool in the Lead-DBS software^8^ for illustrative cases. This was performed based on the recorded targeting coordinates, as well as on the written description of the surgical approach and available figures showing electrode trajectories.

*Statistical analysis*

To date, there are no comparative trials of DBS targets in the same epilepsy population, such that robust comparisons of seizure outcomes across targets were not possible. Instead, we report on the effectiveness of each DBS target in the corresponding patient populations (i.e. mean SR, response rate and seizure freedom). Of note, data was analyzed both at the *study level* (when variables were reported in aggregates for the entire study sample), and at the *patient level* (for the subset of studies that reported information on individual patients). Analyses incorporating variables such as epilepsy type, which are specific to individual patients, were therefore conducted using patient-level data only. Missing data was not imputed and was excluded from analysis.

Additionally, Pearson correlations were used to investigate the relationship between electrode position (x, y and z MNI coordinates) and outcomes for the ANT and CM targets, for which more data points were available. This was done both at the study level and individual patient level. Paired t-tests were also used to compare coordinates in responders vs. non-responders. For the ANT, we also calculated the Euclidean distance to the mmt-ANT junction (as identified in MNI space based on an anatomically accurate representation of the tract) and correlated this variable to outcomes. P-values are reported without correction for multiple comparisons and considered significant below α = 0.05.

*Data availability*

The database resulting from the systematic review process (124 studies, N = 1,210 patients), is available in full as at <https://osf.io/kpwrv/>. The reconstructed electrodes were compiled as a Lead-DBS dataset^8^, which is also available at <https://osf.io/kpwrv/> and can be visualized using the dedicated 3-D viewer.

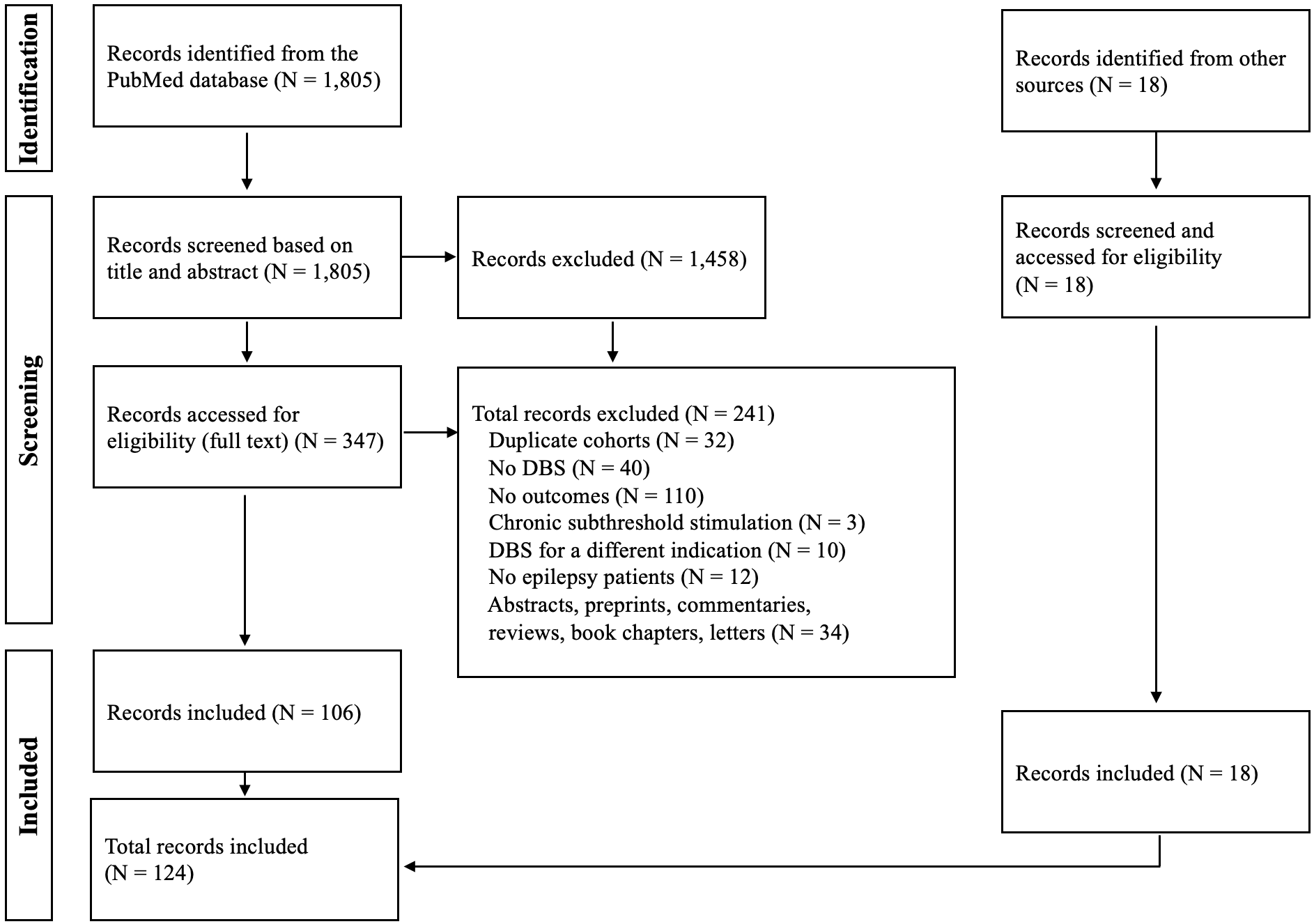

**Fig S1**: **Flow chart of the inclusion and exclusion process.**

**Supplementary results**

*Anterior nucleus of the thalamus*

Sixty studies performed **ANT-DBS, including a total of 801 patients**. Across these studies, the mean SR was 46.3 ± 19.1%, with 54.8% of patients (375/684)^[1]^ classified as responders. Two randomized controlled trials (RCTs) were conducted^9,10,11^. The largest one, the SANTE trial (N = 110) reported a median SR of 40.4% (vs. 14.5% in controls) at the end of the 3 month blinded phase (responder rate: 43%), increasing to 69% at 5 years (open label phase, responder rate: 68%)^9,10^. Herrman et al. (N = 18) found a 22% SR at the end of the 6-month blinded phase, but no significant difference from controls not receiving active stimulation, leading to early termination due to low treatment response^11^.

Patient-level data was available for 412 patients (mean SR: 46.0 ± 55.6% based on N = 281; 251/412 responders; average FU time: 22.4 ± 16.9, range: 1-120 months; Fig. 3). Among these, as expected, **the majority (N = 361) had focal or multifocal epilepsy**. This group had an average SR of 46.1 ± 54.0% (based on N = 238) and included 220 responders (60.9%) and 17 (4.7%) patients achieving seizure freedom at the time of follow-up. Twenty-one patients had generalized epilepsy, with an average SR of 27.0 ± 90.5% (based on N = 17), including 12 responders (57.1%) and one patient (4.8%) achieving seizure freedom. Twelve patients had combined generalized and focal/multifocal epilepsy, with an average SR of 45.7 ± 37.8% (based on N = 12), including six responders (50.0%) and one achieving seizure freedom (8.3%). The remaining 18 patients had unspecified epilepsy types, with an average SR of 68.8 ± 34.6% (based on N =14), including 13 responders (72.2%) and three patients (16.7%) achieving seizure freedom.

Although these findings should be interpreted with caution, and despite the numerically lower SR in patients with generalized epilepsy, there were **no statistically significant differences** in SR outcomes between patients with focal/multifocal, generalized, combined, or unspecified epilepsies (**F(3, 277) = 1.5, p = 0.23).**

Among the patients with focal/multifocal epilepsy, seizure onset zones were located in the temporal lobe for 141 (SR = 44.9% ± 63.7% based on N = 107, 88/141 responders), in the frontal lobe for 53 (SR = 49.9% ± 44.5% based on N = 32 , 32/53 responders), and in the parietal or parieto-occipital region for 10 (SR = 47.4 ± 52.5% based on N = 8, 7/10 responders). Seizure onset lobar localization was unspecified or spanned multiple lobes for the remaining 157 patients. While the SANTE trial suggested that patients with temporal lobe epilepsy may benefit from ANT-DBS the most^12^, here, we found no statistically significant difference in SR between the frontal and temporal lobe groups (t(137) = 0.4, p = 0.68; note that this patient-level analysis does not include the SANTE trial patients, which were only included in our study-level analysis, as no individual-level data was available from the publication).

Finally, it should be noted that three patients received ANT-DBS in the context of status epilepticus. All were considered responders and successfully left the hospital.

Early studies mainly relied on indirect targeting (N = 19 studies), sometimes aided by micro-electrode recordings (MERs). Targeting coordinates were 6.0 mm lateral 8.0 anterior, and 12.0 mm superior to the posterior commissure (PC; N = 3 studies), or **±**4.97 ± 1.07, 1.40 ± 1.86, and 11.15 ± 1.31 mm relative to midcommissural point (MCP; N = 10 studies), corresponding to average MNI coordinates of x = ±5.16 ± 0.97, y = -9.26 ± 3.01, z = 9.35 ± 1.38 mm. More recently, direct targeting has been performed, using specialized MRI sequences capable of visualizing the mammillothalamic tract (mmt) by enhancing myelin signal^13,14^ and aiming for placement of electrodes at the junction between ANT and mmt, which resulted in more anterior MNI coordinates (x = ±5.14 ± 0.81, y = -7.15 ± 1.42, z = 9.94 ± 3.11; N = 17 studies) as compared to studies performing indirect targeting (t(58)=-3.61, p<0.001; with no statistically significant difference in x or z coordinates). Electrodes were implanted using a frontal transventricular (e.g. ^23,249^), extraventricular (e.g. ^15^), or posterior parietal extraventricular approach (e.g. ^16^; see trajectories in Fig. 2).

Leveraging coordinates in our database, we tested two a priori hypotheses from the published literature, the first one being that more anterior electrode placement would be associated with better outcomes (as proposed by Lehtimäki et al.^17^), and the second one being that stimulation of the ANT-mmt junction would be associated with better outcomes (as proposed by Schaper et al. ^15^). Additional analyses using x and z coordinates were also performed and should be considered exploratory.

At the study level (N = 43), more anterior target coordinates were associated with better outcomes (correlation between study-level MNI coordinates and average SR, x: r = 0.29, p = 0.094; y: r = 0.34, p = 0.041; z: r = 0.19, p = 0.29). Additionally, studies with coordinates closer to the ANT-mmt junction reported better outcomes (correlation between Euclidean distance to ANT-mmt junction and average SR: r = -0.55, p < 0.001; Fig. 3).

Individual-level active contact coordinates were available for 5 studies^18–22^ for a total of 52 patients (N = 49 with focal or multifocal epilepsy), including 46 with SR data. Average MNI coordinates for active contacts were x = ±5.12 ± 1.20, y = -6.43 ± 2.30, z = 8.53 ± 2.34. More dorsally located active contacts (z coordinate) were associated with better outcomes (r = 0.37, p = 0.012). There were no significant correlations with x (r = 0.00, p = 0.98) or y coordinates (r = 0.05, p = 0.76). Additionally, responders (N = 35) had more dorsal active contacts than non-responders (N = 17), and there was a trend for more anterior active contacts in this group as well (x = 5.20 ± 0.96 vs. 4.95 ± 0.67, t(50) = 1.00, p = 0.32; y = -6.04 ± 1.91 vs. -7.23 ± 2.50, t(50) = 1.90, p = 0.063; z = 8.99 ± 1.72 vs. 7.57 ± 2.81, t(50) = 2.25, p = 0.028). Finally, as observed at the group-level, active contacts closer to the ANT-mmt junction were associated with better outcomes (r = -0.51, p < 0.001; Fig. 3).

*Centromedian nucleus of the thalamus*

Thirty-two studies performed **CM-DBS, including 208 patients**. The mean SR was 64.2 ± 20.0%, with 136/178 responders (76.4%). Two randomized controlled trials were identified. The ESTEL trial^23^ reported a median SR of 35% at the end of the 3-month blinded phase (57% based on electrographic seizures), with a significantly higher responder rate in the stimulation vs. control group based on electrographic, but not diary-recorded seizures. The study by Fisher et al.^24^ reported a 30% SR at the end of the 3-month blinded on-stimulation phase, which did not significantly differ from the 8% SR observed in the off-stimulation phase.

Patient-level data was available for 178 patients (mean SR: 62.2 ± 37.2% based on N = 168; 133/178 responders; average FU time: 20.7 ± 27.0, range: 0.5-164 months; Fig. 3). As expected, **the majority (N=138) had generalized epilepsy**, with an average SR of 65.3 ± 33.9% (based on N = 132), 108/138 were responders (78.3%), and 7 became seizure-free (5.1%) at follow up. Thirty patients had focal or multifocal epilepsy, with an average SR of 49.6 ± 50.8% (based on N = 27), 19/30 were responders (63.3%), and 1 achieved seizure freedom (3.3%). Ten patients had combined (generalized and focal/multifocal) epilepsy, with an average SR of 54.9 ± 30.5% (based on N = 9), 6/10 were responders (60.0%), and 1 achieved seizure freedom (10.0%).

There was no significant difference in SR between patients with focal/multifocal, generalized, or combined epilepsies (F(2,165) = 2.2, p = 0.11), although numerically higher SR was observed in patients with generalized epilepsy. y.

Notably, 105 patients treated with CM-DBS had Lennox-Gastaut syndrome (LGS) or “LGS-like” syndrome, demonstrating an average SR of 62.3 ± 34.5% (based on N = 103), with 76/105 responders (72.4%). Furthermore, 5 of these patients achieved seizure freedom (4.8%). Seventeen patients treated with CM-DBS had idiopathic generalized epilepsy, with an average SR of 75.2 ± 21.2% (based on N = 16), 16/17 responders (94.1%), and 2 patients achieving seizure freedom (11.8%). Regarding patients with focal or multifocal epilepsies, one study specifically included patients with frontal epilepsies (N=4), with an average SR of 36.0 ± 36.1%.

Additionally, six patients with status epilepticus were treated with CM-DBS and classified as responders. Finally, one patient received CM/Pf-DBS for concomitant epilepsy and Tourette syndrome and became seizure-free at one year. Although the patient’s tics initially improved drastically, the last follow-up revealed only mild improvement according to the Yale Global Tic Severity Scale.

Electrode implantation in the CM (Fig. 2) mainly relied on indirect targeting, which was sometimes complemented by MERs or intraoperative stimulation and EEG recording of recruiting responses. Two studies used short tau inversion recovery MRI to perform direct targeting^25,26^. Average AC-PC target coordinates for CM were **±**9.83 ± 0.86, 0.99 ± 1.55, and 0.61 ± 1.07 mm relative to PC, or **±**8.0 ± 0.89, 10.33 ± 1.37, and 0.67 ± 0.52 mm relative to MCP. This corresponded to the following average MNI coordinates: x= ±9.51 ± 1.56, y= -21.79 ± 1.58, z =-1.79 ± 0.72.

Given the reliance on direct targeting, there was limited variance in targeting coordinates, such that no significant correlations could be identified between coordinates and SR at the study level (N = 26 studies). Patient-level active contact coordinates were only available for two studies^27,28^ (N = 19 and 14), with only the study by Warren et al. reporting corresponding percent SR. Across these studies, average MNI coordinates for active contacts were x = ±10.02 ± 0.79, y = -19.63 ± 1.37, z = 0.82 ± 1.18. An additional two studies by Velasco et al.^29,30^ represented the electrode positions of responders vs. non responders (note that responders were defined as having a SR >80% in these studies; total N = 26) but precise coordinates could not be extracted due to issues with matching the figure axes. This data is visualized in Fig. 4. Overall, available data suggests that greater SR is associated with stimulation of the anterior and inferolateral (parvocellular) part of CM in patients with generalized epilepsy (with significantly lower active contacts in responders vs. non responders in our analysis; t(40) = -2.46, p = 0.018, 0.46 ± 1.58 vs. 1.55 ± 1.17; and no significant difference on the other axes). The data from Son et al.^28^ suggests that better SR may be associated with stimulation of the MD nucleus in patients with multifocal epilepsy (with significantly higher active contacts in responders vs. non responders in our analysis; t(17) = 2.75, p = 0.014, 3.33 ± 2.05 vs. -0.06 ± 3.19; and no significant difference on the other axes). These results should be interpreted with caution given the small sample size.

*Hippocampus, amygdala and parahippocampal cortex*

Fifteen studies performed **hippocampal DBS, including a total of 89 patients**. The mean SR was 64.5 ± 19.1%, with 68/87 patients (78.2%) classified as responders. Randomized controlled trials included Velasco et al. (2007)^31^, reporting a median SR of 72% over 18 months (open label phase; no statistics reported for the 1-month blinded phase); Cukiert et al. (2017)^32^, with a median SR of 66% at the end of the 6-month blinded phase (demonstrating superiority to sham stimulation); and Tellez-Zenteno et al. (2006)^33^, showing a 15% median SR in the blinded ON as compared to the blinded OFF stimulation phase (non-significant).

Patient-level data was available for 87 patients (average FU time: 35.2 ± 20.9, range: 3-96 months), **all with focal or multifocal temporal lobe epilepsy** (Fig. 3). In most cases, these patients received DBS because they were not considered good candidates for resection^33,34,35,36,37^. The mean SR in this group was 65.2 ± 34.5% (based on N = 85), with 68/87 patients (78.2%) classified as responders and 14 patients (16.1%) becoming seizure free. Specifically, this group included 36 patients with bilateral and 51 patients with unilateral temporal lobe seizure onset. Average SR was 75.9 ± 25.7 in patients with bilateral (32/36 responders, i.e. 88.9%, 10 seizure-free), and 58.2 ± 37.9% (36/51 responders, i.e. 70.6%, 4 seizure-free) in patients with unilateral onset, a difference that reached statistical significance (t(83) = 2.4, p = 0.019), although this result should be interpreted with caution. Additionally, although previous studies suggested that patients with hippocampal sclerosis may demonstrate lower response to DBS of the hippocampus^31,38^, we found no significant difference in outcomes between patients with mesial temporal lobe sclerosis (N = 52, SR = 63.8 ± 36.7%, 40 responders, i.e. 76.9%) and patients with non-lesional imaging (N = 29, SR = 65.6 ± 32.8%, 22 responders, i.e. 75.9%; t(78) = -0.22, p = 0.83).

Finally, one patient received DBS of the posterior sylvian junction after the hippocampal DBS electrode migrated to this location one month after surgery. Despite this, the patient was classified as a responder.

There was considerable variability in targeting strategies. Most studies used a parasagittal occipital approach to implant electrodes along the main axis of the hippocampus (as directly visualized on MRI), with only one study using a frontal approach^39^, placing the deepest contacts in the parahippocampal gyrus. While most studies positioned the most distal contact in the hippocampus head, different electrode models were used (with electrode span up to 34.5 mm), which resulted in more or less extensive coverage of the hippocampus (Fig. 2). Variability in stimulation parameters also contributed to this, with some studies activating all four electrode contacts and others activating only specific ones. Two studies aimed to place the stimulating contacts as close as possible to the seizure onset zone location as identified with stereo-EEG^40,41^. Seven studies implanted electrodes bilaterally, five unilaterally, and three included both depending on seizure onset laterality. In summary, different subdomains of the hippocampus may have been stimulated across studies, and whether this contributed to differences in outcomes is unclear. Additionally, one study implanted an additional electrode in the amygdala^42^, and one targeted the parahippocampal cortex to avoid stimulating the hippocampus when hippocampal sclerosis was present^43^.

*Motor targets*

Fifteen studies performed DBS targeting motor regions, including **2 studies targeting the caudal zona incerta (cZi, N = 5), 9 studies targeting the subthalamic nucleus, 3 studies targeting the STN / substantia nigra pars reticulata region (STN and STN/SNr: N = 44),** and **one study targeting the ventral intermediate nucleus (Vim, N = 1).**

Patient-level data was available for all **50 patients**, with a mean SR of **55.6 ± 70.3% (based on N = 43),** including **38/50 responders (76.0%)** and **eight patients (16.0%)** achieving seizure freedom at the time of follow up (average FU time: 32.00 ± 25.7, range: 1-108 months; Fig. 3).

Of these, 27 patients were reported as having motor seizures and/or seizures originating from central regions, with an average SR of 69.3 **±** 42.3% (based on N = 26), corresponding to 23/27 responders (85.2%) and 5 patients (18.5%) achieving seizure freedom. Another 10 patients had progressive myoclonic epilepsy, including 8/10 responders (80.0%) and two patients (20.0%) achieving seizure freedom. Percent SR was only reported for 4 of these patients (average: 75.8 **±** 31.8%). One patient had juvenile myoclonic epilepsy and improved by 87.5%. The remaining 12 patients, with diverse epilepsy types and lobar localizations, had an average SR of 16.6 **±** 110.5% (based on N = 12; 6/12 responders, i.e. 50%).

For the cZI, while targeting coordinates were similar across the two studies, one used a frontal approach, and the other one a parieto-occipital approach (**±**13, -6, -2 relative to MCP), which may have resulted in different structures being stimulated. The Vim implantation used indirect targeting with micro-electrode recordings and scalp EEG. In this patient, the initial target was the CM, but the electrode was implanted in the Vim after serendipitous observation of recruiting responses and modulation of epileptic activity with stimulation of this nucleus during surgery^44^.

Early suggestion that the optimal target for epilepsy in the subthalamic region is lower and medial than the optimal STN target for Parkinson’s Disease initiated a debate on whether the STN or SNr should be targeted for epilepsy^45,46^. As a consequence, some studies implanted electrodes in the dorsolateral STN^47,48^, while others placed one or more of the four contacts in the SNr^46,49,50^ (Fig. 2). Additionally, different electrode models were used, adding to the variability of final stimulation sites and clouding the relationship between target location and outcomes. Both indirect targeting with MERs and direct targeting were used across studies. Average AC-PC coordinates were **±11.15** ± 1.06, -3.11 ± 0.99, and -5.06 ± 1.20 mm relative to MCP, corresponding to average MNI coordinates of x = ±11.42 ± 1.01, y = -14.54 ± 1.23, z = -8.40 ± 1.29 (N = 10 studies).

*Multitarget DBS (ANT+)*

Six studies performed ANT-DBS alongside other targets, implanting two pairs of electrodes. Secondary targets included the **CM (N = 14 patients), NAc (N = 3), Vim (N = 1), hippocampus (N = 4), pulvinar (N = 2), mediodorsal nucleus (N = 2), and primary motor cortex - supplementary motor area (N = 1),** for a total of **26 patients.**

Patient-level data was available for all **26 patients with a mean SR of 52.0 ± 38.8% (based on N = 26), 16/26 responders (61.5%) and no patient achieving seizure freedom** (average FU time: 45.5 ± 36.3, range: 3-100 months). Rationales for these dual target implantations included combining an approved target (the ANT) with the more novel target being explored^51^, implanting additional electrodes in the seizure onset zone^52^ or combining targets to treat patients who don’t correspond to the populations typically treated with ANT- or CM-DBS, and whose seizures may be **difficult to treat** with one of these targets alone^53,54^. For example, in the study by Yang et al.,^55^ the CM target was added for patients with generalized or multifocal non-limbic seizures, the pulvinar for posterior quadrant epilepsy, the mediodorsal nucleus for motor semiology, and the STN for myoclonic seizures or seizures originating from the supplementary motor area. The most common target combination was ANT- and CM-DBS, with 15 patients, including 10 with a combined epilepsy type. Patients with ANT and CM-DBS experienced a SR of 58.4 **±** 35.1% (based on N = 15; 10/15 responders, i.e. 66%, no patient achieved seizure freedom).

While surgical targeting for multitarget implantations typically followed standard procedure for the corresponding structures, Yang et al.^55^ used eight-contact electrodes and adjusted trajectories in order to target two thalamic nuclei with a single electrode, including the ANT and MD, and the pulvinar and CM (Fig. 2).

*Other limbic targets*

Four studies investigated deep brain stimulation targeting limbic regions, including two studies targeting the **nucleus accumbens (N = 9 patients),** one study targeting the **mammillothalamic tract (N = 2)**, and one study targeting the **fornicodorsocommissural tract (N = 4)**. In the randomized controlled trial by Kowski et al. (2015)^56^ investigating DBS of the NAc, 3 patients were responders during the 3-month blinded ON stimulation phase, as compared to none in the OFF phase. A single-blinded controlled trial of low-frequency fornicodorsocommissural tract stimulation by Koubeissi et al. (2022)^57^ demonstrated a median reduction of **68%** over **8 months, with significantly lower seizure frequency in the ON vs. OFF single-blinded stimulation phase**.

Patient-level data was available for all 15 patients, including **9/15 responders (60.0%)** and **2 patients (13.3%)** achieving seizure freedom at the time of follow-up (average FU time: 9.5 ± 6.8, range: 3-24 months). Mean SR was **52.7 ± 69.4%**, with missing data for 3 non-responders. Of the nine patients who received nucleus accumbens stimulation, all had **focal or multifocal epilepsy** originating from the **frontal (N = 4)** and/or **temporal lobes (N = 6),** with **5/9 responders (55.6%)** but no patients achieving seizure freedom. Missing data for 3 of the 4 non-responders prevented robust calculation of average SR for these patients. Among the two patients who underwent mammillothalamic tract stimulation, both with **hypothalamic hamartomas**, **one** achieved seizure freedom and the other experienced a SR of 84%. The remaining four patients, all with **bilateral mesial temporal lobe epilepsy,** received stimulation of fornicodorsocommissural tract, which connects both hippocampi, achieving a mean SR of **64.0 ± 36.0%,** with **2 responders** and **one patient** achieving seizure freedom.

Targeting relied on direct visualization of the target structures. Additionally, the fornicodorsocommissural target was confirmed by intraoperative recording of stimulation-induced hippocampal responses. Electrode trajectories are shown in Fig. 2.

*Hypothalamus*

Five studies performed **hypothalamic DBS, including a total of 12 patients**. Patient-level data was available for all 12 patients. There were **8/12 responders (66.7%)** and one patient achieving seizure freedom (8.3%). Percent SR was only available for the 8 responders, and not for the 4 non-responders, with an average of 87.8 **± 17.9%** (average FU time: 36.1 ± 23.2, range: 2-82 months). In 11 patients, the hypothalamus was chosen as a target because of **comorbid intractable aggressive behavior** (8 responders, 3 non-responders), including 5 patients with generalized, 3 with focal or multifocal, and 2 with combined generalized and focal epilepsy. Of note, all studies reported improvements in comorbid aggressive behavior. The remaining patient had focal epilepsy associated with a **hypothalamic hamartoma** and was a non-responder.

Among the four studies reporting targeting coordinates, three used the same target (**±**2, -3, -5 mm relative to MCP for the tip), with the other using slightly different coordinates (**±**3.5, 0, -2 mm relative to MCP, Fig. 2). Target was confirmed with MERs and stimulation to assess physiological responses. The last study aimed to insert electrodes directly within the hamartoma, such that template space coordinates couldn’t be obtained.

*Visual Targets*

Three studies performed DBS on targets pertaining to the visual system, including a total of **three patients (two pulvinar, one lateral geniculate nucleus)**. The mean SR was **75.6 ± 19.2% (based on N = 3),** with all patients being diagnosed with focal or multifocal epilepsy and being classified as responders, though none achieved seizure freedom at the time of follow up (average FU time: 14.0 ± 9.5, range: 5-24 months). Two patients had **occipital lobe epilepsy** (one with pulvinar, one with lateral geniculate nucleus DBS). The last patient had “**temporal plus” epilepsy** and was targeted in the pulvinar, on the basis of extensive connections of this nucleus with parietal, temporal and occipital cortices^58^.

The lateral geniculate nucleus was targeted with the help of tractography, MERs of responses to light flashes, and macrostimulation. There was no detail on the pulvinar implantations (Fig. 2).

*Cerebellum*

Finally, one study performed **cerebellar DBS** **in five patients**, resulting in an average SR of **71.6 ± 13.3%**, with all patients classified as responders. Patients were selected based on having **intractable motor seizures**^59^. Based on patient-level data, **4** had focal or multifocal epilepsy with an average SR of **71.0 ± 15.3%**, and all were responders, though none achieved seizure freedom. The remaining patient had generalized epilepsy with a SR of **72.7%**. Average FU time was 19.6 ± 6.2 (range: 11-24) months.

The superomedial cerebellar cortex was targeted with four-button electrodes, and stimulation was delivered through all four contacts, resulting in the stimulation of a large area of the upper medial surface (Fig. 2).

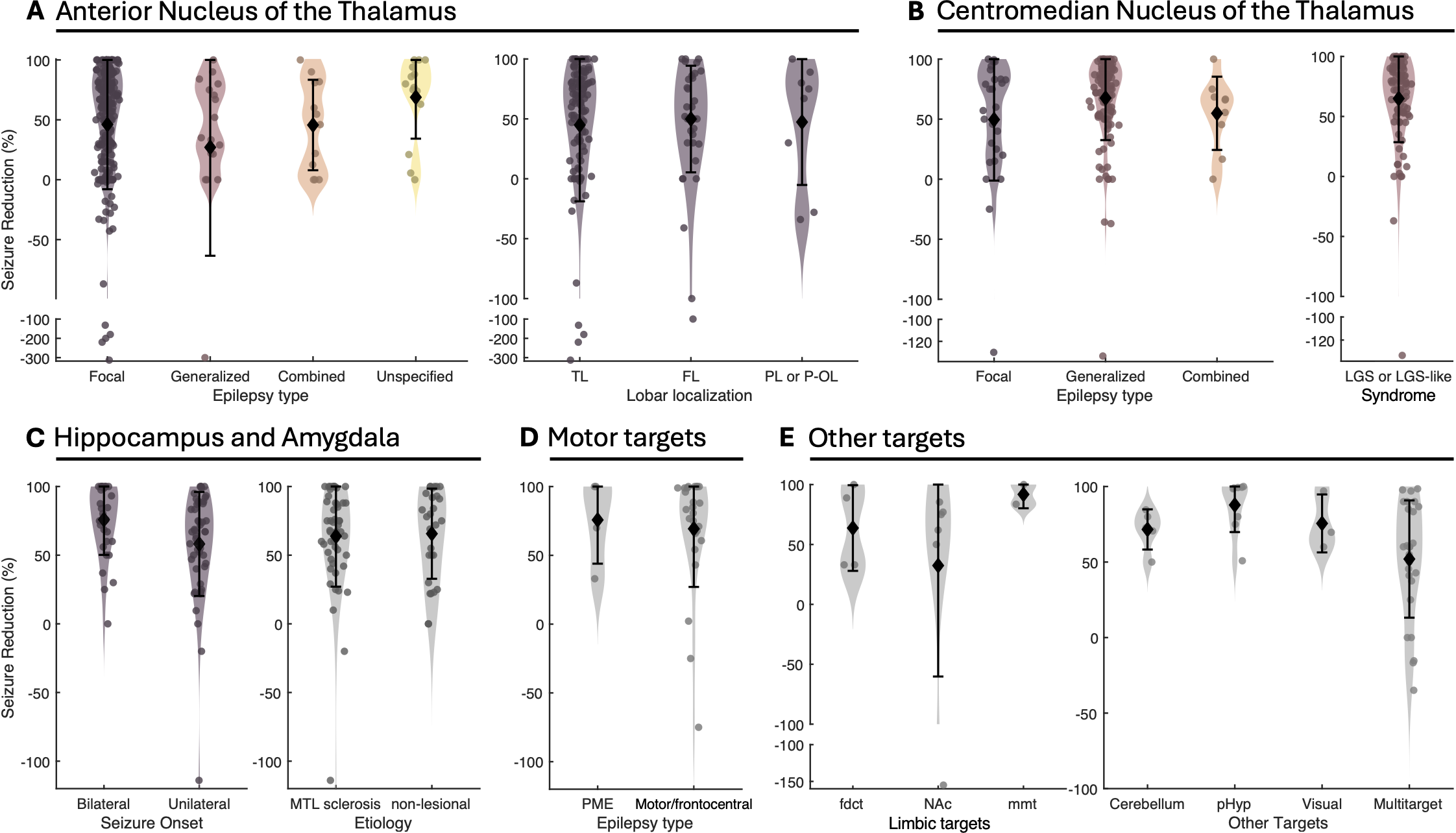

**Fig S2: Seizure reduction across DBS targets and epilepsy types.** Overall, while average seizure reduction was above 50% for many groups and targets, seizure freedom was rarely achieved. (A) For ANT-DBS, seizure reduction was numerically smaller for patients with generalized epilepsy, although this difference did not reach statistical significance. Among patients with focal or multifocal epilepsy, there was no evident difference in outcomes depending on seizure onset lobar localization (TL: temporal lobe, FL: frontal lobe, PL or P-OL: parietal or parieto-occipital). (B) For CM-DBS, there was a tendency for patients with generalized epilepsy to experience larger seizure reductions. Among the patients with generalized epilepsy, many patients had LGS or “LGS-like” syndrome. (C) For DBS of the hippocampus and amygdala in patients with temporal lobe epilepsy, patients with bilateral seizure onset experienced significantly higher seizure reduction than patients with unilateral seizure onset. We found no significant difference between patients with mesial temporal lobe sclerosis and patients with non-lesional imaging. (D) DBS of motor targets (STN, SNr, Vim) led to important seizure reductions in patients with motor seizures and/or seizure onset localized to central regions as well as in patients with progressive myoclonic epilepsy (PME). (E) Outcomes are represented for less common targets such as the fornicodorsocommissural tract (fdct), NAc, mammillothalamic tract (mmt), cerebellum, posterior hypothalamus (pHyp), targets pertaining to the visual system (lateral geniculate or pulvinar), as well as for multitarget DBS (see main text for detail). Note the broken axes in panels A, B and E.

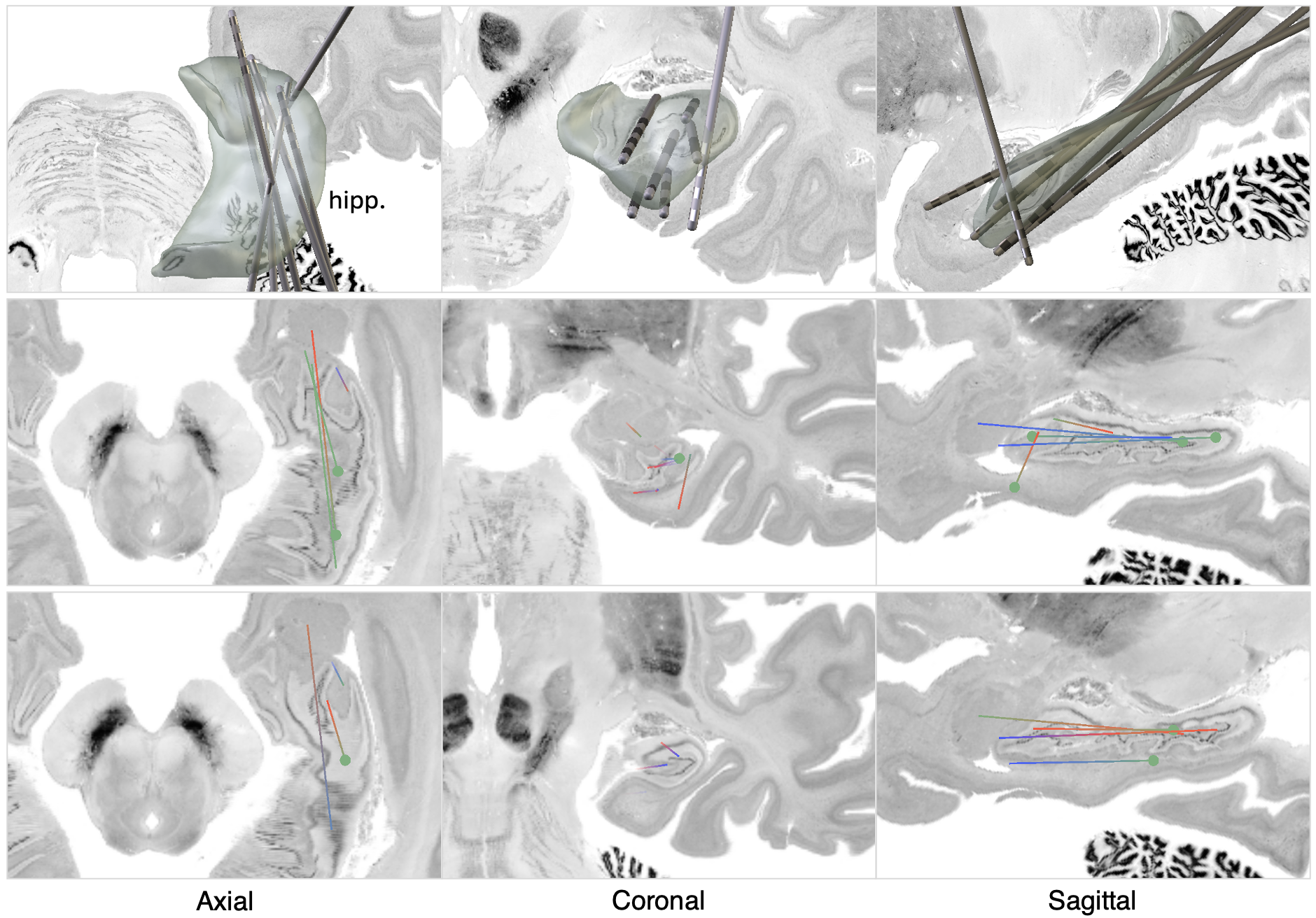

**Figure S3: Electrode positions for DBS studies targeting the mesial temporal lobe** (hippocampus, amygdala and parahippocampal cortex), 3D (top row) and 2D (bottom rows) views overlaid on the Big Brain template ^60,61^. For the 2D panels, each line represents an electrode, and the MNI template was rotated by 37° in the anterior-posterior direction to show the main axis of the hippocampus in the axial plane.

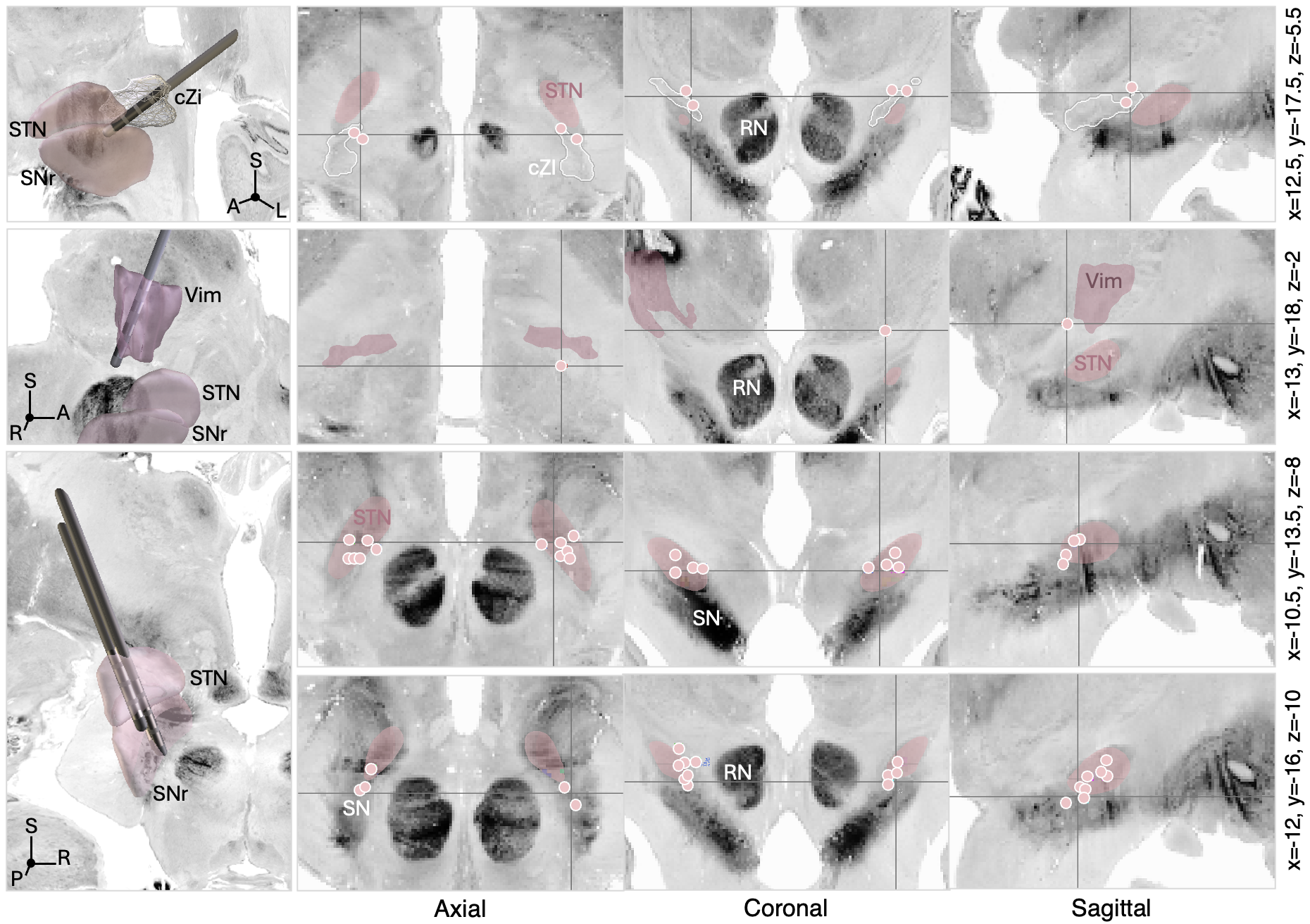

**Figure S4: Electrode positions and targeting coordinates for DBS studies investigating motor targets** (in order, caudal zona incerta, ventral intermediate nucleus of the thalamus, and subthalamic nucleus / substantia nigra pars reticulata). 3D and 2D views are overlaid on the Big Brain template^60,61^, and anatomical structures are shown as defined in the DISTAL^62^ and CIT168 atlases^63^. Electrode coordinates were projected to common planes for visualization using the maximal intensity projection feature of the FSLeyes software. Coordinates on the right indicate the position of the location cursor.

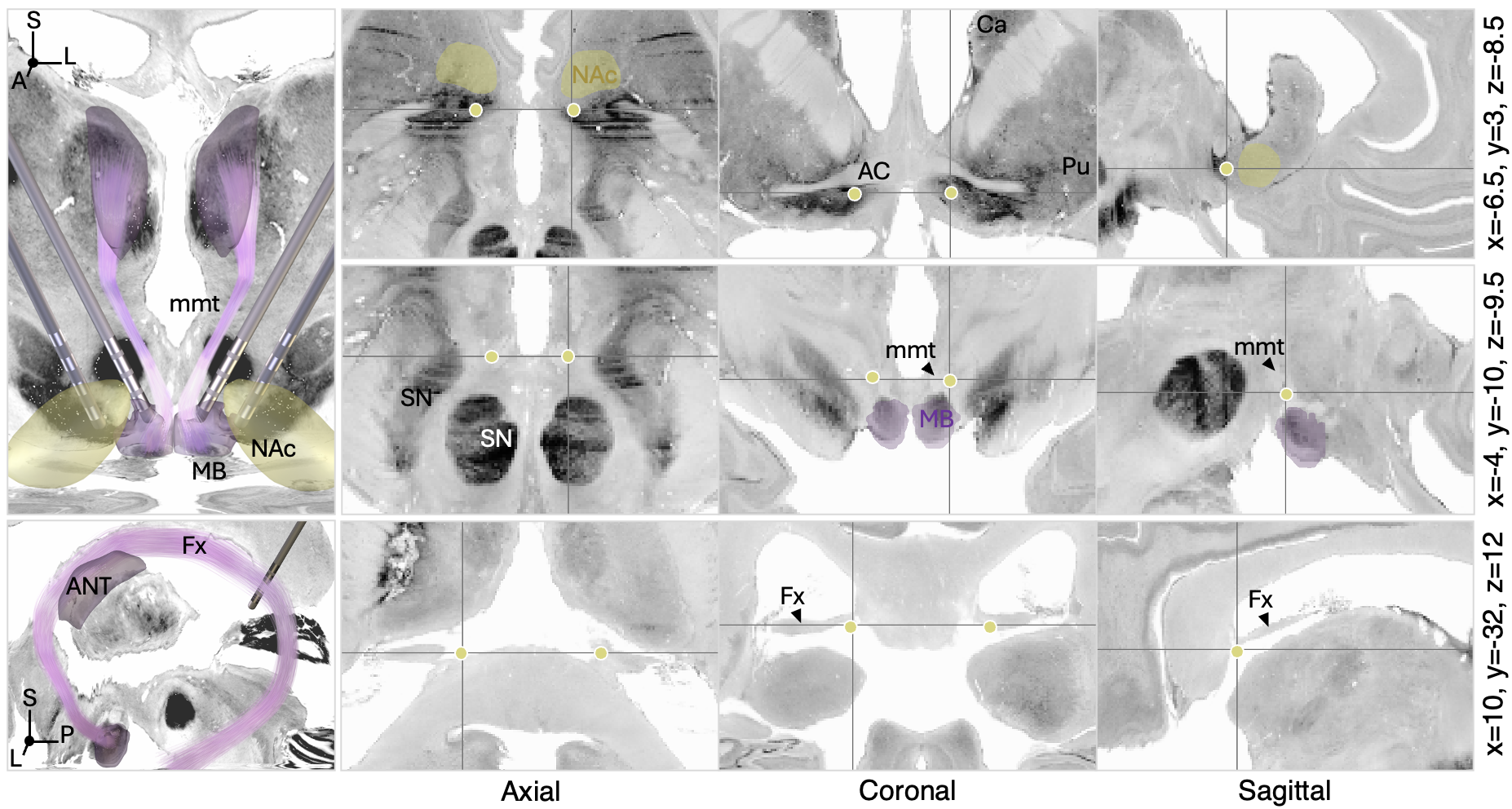

**Figure S5: Electrode positions and targeting coordinates for DBS studies investigating other limbic targets** (in order, nucleus accumbens, mammillothalamic tract, and fornix). 3D and 2D views are overlaid on the Big Brain template^60,61^, and anatomical structures are shown as defined in the Morel^64^ and CIT168 atlases^63^, as well as in the Atlas of the human hypothalamus^65^. Electrode coordinates were projected to common planes for visualization using the maximal intensity projection feature of the FSLeyes software. Coordinates on the right indicate the position of the location cursor.

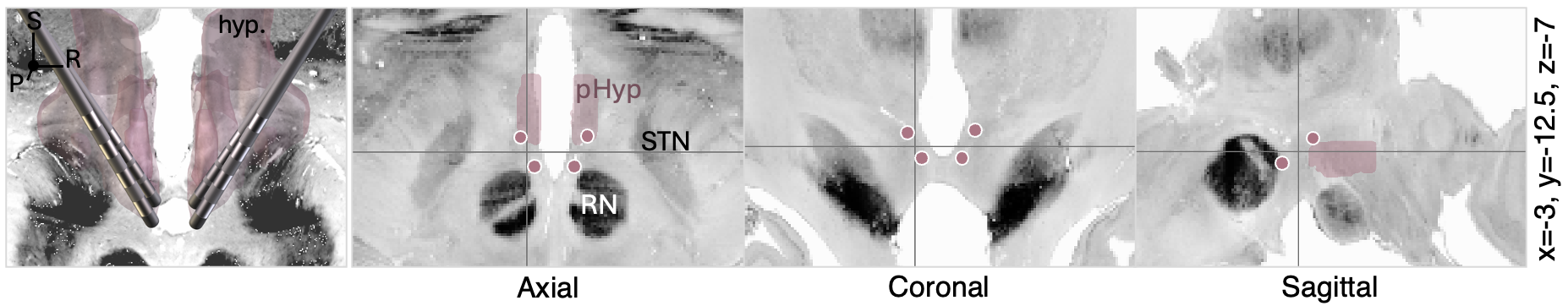

**Figure S6: Electrode positions and targeting coordinates for DBS studies targeting the hypothalamus**. 3D and 2D views are overlaid on the Big Brain template^60,61^, and the posterior hypothalamic nucleus is shown as defined in the the Atlas of the human hypothalamus^65^. Electrode coordinates were projected to common planes for visualization using the maximal intensity projection feature of the FSLeyes software. Coordinates on the right indicate the position of the location cursor.

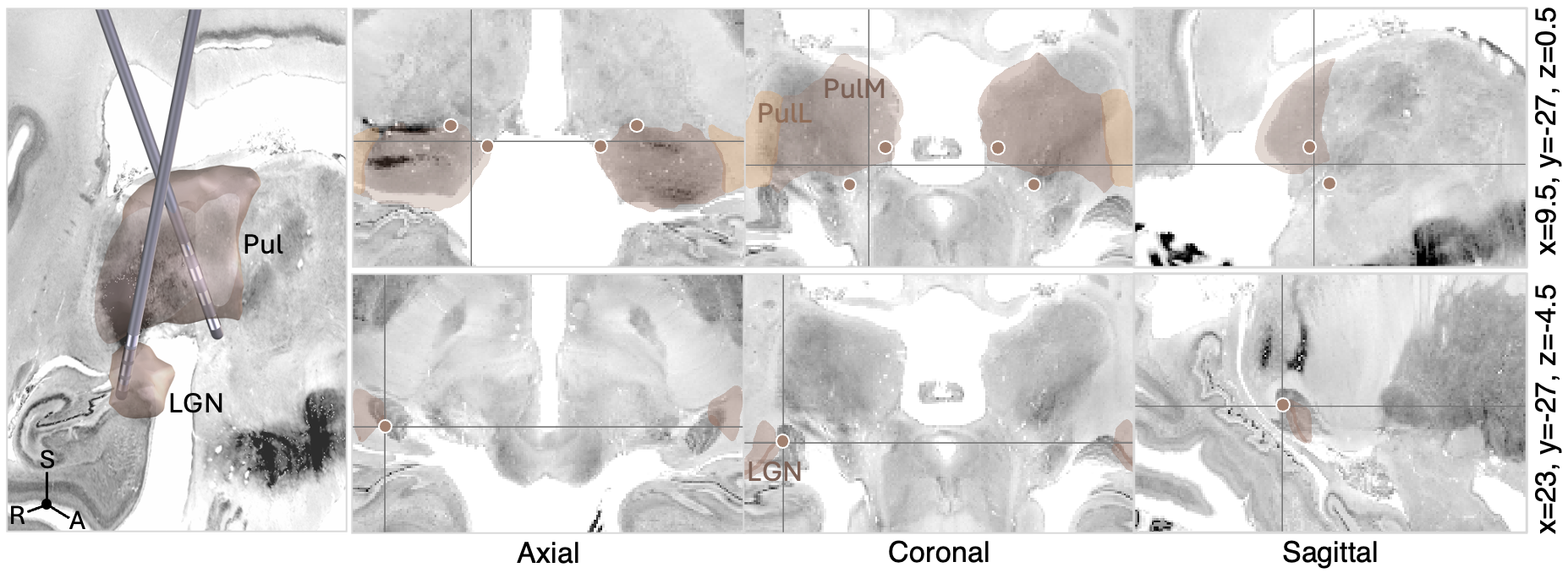
**Figure S7: Electrode positions and targeting coordinates for DBS studies investigating visual targets** (in order, pulvinar and lateral geniculate nucleus). 3D and 2D views are overlaid on the Big Brain template^60,61^, and anatomical structures are shown as defined in the Morel atlas^64^. Electrode coordinates were projected to common planes for visualization using the maximal intensity projection feature of the FSLeyes software. Coordinates on the right indicate the position of the location cursor.

| Study (N) | Method | Main findings |
| --- | --- | --- |
| *ANT-DBS* |  |  |
| Krishna et al., 2016^66^ N=16 | Stimulation heat maps based on stimulation volumes | Stimulation hotspot for responders was located in the anteroventral part of the ANT, immediately superior and posterior to the ANT-mmt junction |
| Lehtimaki et al., 2016^17^ N = 15 | Responding and non-responding contact coordinates (AC-PC) | Responding contacts were located more superior and anterior in AC-PC space, and in the anterior aspect of ANT |
| Koeppen et al., 2018^67^ N = 10 | Target and active contact coordinates (AC-PC) and stimulation volume overlaps | Higher electrode placement and active contacts were associated with better SR |
| Middlebrooks et al., 2018^68^ N = 6 | Normative functional connectivity profiles from stimulation volumes | Responders had greater connectivity to the DMN and consistent anticorrelation with the hippocampus which was not seen in non-responders |
| Järvenpää et al., 2020^69^ N = 27 | Qualitative analysis of active contact locations | Patients with contacts within the ANT or at its anterolateral border had higher SR than patients with contacts outside the ANT |
| Schaper et al., 2020^70^ N = 20 | Active contact coordinates (AC-PC) and distance to mmt junction | Distance from active contacts to the ANT-mmt junction correlated with SR |
|  | Stimulation heat maps based on stimulation volumes | Stimulation hotspot was located at the ANT-mmt junction for responders and was less localized for non-responders |
| Costa-Gertrudes et al., 2021^71^ N = 14 | Qualitative analysis of active contact coordinates (MNI space) | Proposed an optimal target anterior and inferior in the ANT, slightly medial to the ANT-mmt junction |
| Gross et al., 2021^9^ N = 101 | Qualitative assessment of electrode placement | Patients with "good" bilateral placement (i.e., trajectories passing through the target region within the ANT and superior to the mmt) had a significantly higher probability to be responders |
| Guo et al., 2021^72^ N = 19 | Active contact coordinates (MNI space) | Active contacts were more dorsal in responders and more dorsal active contacts were associated with higher SR |
| Freund et al., 2023^73^ N = 7 | Active contact coordinates (AC-PC) and distance to mmt junction | Active contacts were closer to the ANT-mmt junction in responders than in non-responders, and distance from active contacts to the ANT-mmt junction correlated with SR |
| Schaper et al., 2023^74^ N = 30 | Functional connectivity to an a-priori network (normative connectome) | SR was associated with functional connectivity of stimulation sites to a specific brain network derived from lesions causing epilepsy and defined by functional connectivity to the basal ganglia and cerebellum |
| Xu et al., 2023^75^ N = 18 | Normative functional connectivity profiles from stimulation volumes | Greater functional connectivity between stimulation volumes and seizure foci was associated with better outcomes |

| *CM-DBS* |  |  |
| --- | --- | --- |
| Velasco et al., 2000^29^ N = 13 | Qualitative analysis of electrode position (AC-PC) | Patients with good outcomes (>80% SR) had electrode placements in the ventrolateral part of the PC (parvocellular), and patients with worse outcomes had more dorsal, posterior or medial placements. |
| Velasco et al., 2006^76^ N = 13 | Qualitative analysis of electrode position (AC-PC) | Patients with good outcomes (>80% SR) had electrode placements grouped in the anterolateral part of the CM, while electrodes were placed in more posterior, superior and medial regions of CM in patients with worse outcomes |
| Son et al., 2016^77^ N = 14 | Active contact coordinates (AC-PC) | No significant correlation between coordinates and outcomes. Patients with multilobar epilepsy had significantly higher active contact coordinates than patients with generalized epilepsy |
| Warren et al., 2020^78^ N = 20 | Probabilistic stimulation mapping (sweet-spots) | Stimulation of the anterior and inferolateral "parvocellular" CM border (extending to the ventral lateral nucleus) was associated with greater SR |
|  | Structural connectivity profiles (normative and disease-matched connectomes) | Structural connectivity from stimulation sites to areas of EEG-fMRI activation previously found to express epileptic activity in LGS was associated with greater SR |
| Torres Dias et al., 2021^79^ N = 10 | Distance from stimulation volumes to CM | Better SR was associated with shorter distance between the stimulation volume and CM |
|  | Structural and functional connectivity profiles (disease-matched and normative connectomes) | Greater SR was associated with connectivity from stimulation sites to a reticular system network encompassing sensorimotor and supplementary motor cortices as well as the brainstem and cerebellum |
| Yang et al., 2022^80^ N = 13 | Heat map created based on active contact positions (MNI space) and corresponding outcomes | Stimulation of the anteromedial region of the CM was associated with greater SR |
| *Hippocampus-DBS* |  |  |
| Bondallaz et al., 2013^35^ N = 8 | Qualitative analysis of active contact locations | Active contacts were closer to the subiculum in responders than non-responders |
| Cukiert et al., 2020^81^ N = 25 | Qualitative analysis of active contact locations | No relationship between active contact location in specific sectors of the hippocampus (central, superior, inferior, media, lateral) and outcomes |

**Table S1: Summary of studies investigating the relationship between electrode placement and treatment outcomes.** Studies using stimulation mapping, contact location analysis, or connectivity-based approaches were considered. Please note that this list may not be exhaustive.

| # | Trajectory ID  Electrode model | Target | Coordinates | References |
| --- | --- | --- | --- | --- |
| 1 | Ant1SANTE  Medtronic 3387 | Ant  Transventricular indirect targeting | -4.4, -7.3, 10.1;  4.6, -7.5, 9.9 | 9;82 |
| 2 | Ant2Hodaie2002  Medtronic 3387 | Ant Transventricular indirect targeting | -5.9, 13.7, 10.5;  5.9, -13.9, 10.3 | 83;84 |
| 3 | Ant3transventriculardirect  Medtronic 3389 | Ant Transventricular direct targeting (landmark: mmt) | -4.5, -8.1, 7.2;  5.2, -9.3, 8.2 | 72;71;85;73 |
| 4 | Ant4Posteriorparietal extraventricular  Medtronic 3389 | Ant  Direct targeting, posterior parietal extraventricular approach (landmark: mmt) | -4.5, -8, 14;  6, -8, 15 | 86;68;87;88 |
| 5 | Ant5Extraventricular  Medtronic 3389 | Ant  Extraventricular frontal approach, direct targeting (landmark: mmt) | -6.2, -7.6, 7.2;  6.4, -7.9, 6.9 | 70 |
| 6 | Cm1Velasco2006  Medtronic 3389 | Cm  Indirect targeting | -10.4, -22.9, 2.1;  9.9, -23.2, -2.3 | 76;89;90;91;80;92;93;94;95 |
| 7 | Hipp1Cukiert2020  Medtronic 3391 | Hippocampus | -30.1, -12.4, 22.6;  31.1, -12.4, -22.5 | 81;96 |
| 8 | Hipp2SaucedoAlvarado 2022  Medtronic 3391 | Parahippocampal cortex | -22.5, -12.6, -29.5;  21.9, -12.6, -29.6 | 97 |
| 9 | Hipp3Jin2016  Medtronic 3387 | Hippocampus  Frontal approach | -32, -12.8, -21.3,  31.5, -11.8, -21.5 | 37 |
| 10 | Hipp4Sobstyl2024  Medtronic 3387 | Hippocampus | -26.2, -11.8, -29.9 | 34 |
| 11 | Hipp5Wang2021  PINS L303 | Hippocampus | -24.8, -8.8, -27.2;  23.8, -8.1, -26.4 | 98 |
| 12 | HippAmy1Boon2007  Medtronic 3387 | Amygdala | -21.3, -1.3, -23.4;  21.29, -0.31, -23.4 | 99 |
| 13 | HippAmy2Boon2007  Medtronic 3387 | Hippocampus | -29.3, -13.4,-18.2;  29.3, -12.4, -18.2 | 99 |
| 14 | StnSnr1Wille2011  Medtronic 3389 | STN/SNr | -11.1, -14.9, -9.1;  11.1, -15.4, -9.2 | 100 |
| 15 | StnSnr2Shan2024  Medtronic 3389 | STN | -11.5, -13.5, -7.4;  10.4, -13.2, -7.7 | 101;102 |
| 16 | StnSnr3diGiacopo2019  Medtronic 3387 | STN/SNr | -11.2, -15.0, -9.1;  11.1, -15.4, -9.3 | 103 |
| 17 | Vim1VilelaFilho2023b  Medtronic 3387 | cZI | -13.3, 16.7, -4.6; 13.1, -17.1, -4.9 | 104 |
| 18 | cZI1Franzini2008  Medtronic 3389 | Vim | -13, -18, -2 | 105 |
| 19 | AntMd1Yang2024  Boston scientific Vercise | Ant/MD | -3.1, -16.9, 2.2;  3.4, -15.4, 3.4 | 106 |
| 20 | CmPul1Yang2024  Boston scientific Vercise | Cm/Pul | -10, -20.9, 3.25;  9.9, -20.5, 3.25 | 106 |
| 21 | NAc1Schmitt2014  Medtronic 3387 | NAc | -6.8, 3, -8.5;  7.5, 2.7, -8.7 | 107;56 |
| 22 | fdct1Koubeissi2022  Medtronic 3389 | Fornico-dorsocommisural tract | -10.4, -32, 11.7;  10.4, -32, 11.7 | 57 |
| 23 | mmt1Khan2009  Medtronic 3389 | mmt | -4, -10, -9.5; 4, -10, -9.1 | 108 |
| 24 | pHyp1Franzini2008  Medtronic 3389 | pHyp | -2.1, -14.2, -8.4;  2.1, -14.3, -8.4 | 105;109 |
| 25 | pHyp2Gouveia2021  Medtronic 3387 | pHyp | -2.1, -14.2, -8.4;  2.1, -14.3, -8.4 | 110 |
| 26 | pHyp3Torres2012  Medtronic 3389 | pHyp | -3.6, -10.9, -5.4;  3.7, -11.1, -5.5 | 111 |
| 27 | LGN1VilelaFilho2023  Medtronic 3389 | LGN | 23, -27.0, -4.4 | 112 |
| 28 | Pul1Yan2023  Medtronic 3387 | Pul | -12, -25, -2;  12, -25, -2 | 113 |
| 29 | NA  Resume 4-button electrodes | Cerebellum, superomedial surface | -9.5, -57.0, -6.3;  9.5, -57.0, -6.3 | 59 |

**Table S2: Summary of trajectories used for each target**

|  | Available at study level | Available at patient level |
| --- | --- | --- |
| Epilepsy syndrome | NA | 468/789 |
| Epilepsy type | NA | 763/789 |
| Etiology | NA | 481/789 |
| Age | 119/124 (average) | 746/789 |
| Sex | 111/124 (ratio) | 624/789 |
| Follow up time | 117/124 (average) | 752/789 |
| Seizure reduction | 100/124 (average) | 631/789 |
| Responder status | 118/124 (ratio) | 787/789 |
| Electrode coordinates | 92/124 | 90/789 |

**Table S3: Summary of available data at the study and patient level**, representing the number of studies or patients for which each variable was reported in the corresponding publication.

**List of included studies**

Agashe S, et al. Centromedian Nucleus of the Thalamus Deep Brain Stimulation for Genetic Generalized Epilepsy: A Case Report and Review of Literature. Front Hum Neurosci. 2022;16:858413.

Alanazi FI. Modulation of neuronal activity in human centromedian nucleus during an auditory attention and working memory task. 2024.

Alcala-Zermeno JL, et al. Centromedian thalamic nucleus with or without anterior thalamic nucleus deep brain stimulation for epilepsy in children and adults: A retrospective case series. Seizure. 2021;84:101–107.

Alcala-Zermeno JL. Invasive neuromodulation for epilepsy: Comparison of multiple approaches from a single center. 2022.

Andrade DM. Dravet syndrome and deep brain stimulation: Seizure control after 10 years of treatment.

Anderson DG, Ne AH, Miller J, Krause A. Deep Brain Stimulation in Three Related Cases of North Sea Progressive Myoclonic Epilepsy from South Africa.

Bonda D. Deep Brain Stimulation of Bilateral Centromedian Thalamic Nuclei in Pediatric Patients with Lennox-Gastaut Syndrome: An Institutional Experience. World Neurosurg. 2024.

Bóné B, et al. Pregnancy and deep brain stimulation therapy for epilepsy. Epileptic Disord. 2021;23.

Buenzli JC, et al. Deep brain stimulation of the anterior nucleus of the thalamus increases slow wave activity in non-rapid eye movement sleep.

Cukiert A, Cukiert CM, Burattini JA, Mariani PP. Hippocampal deep brain stimulation: a therapeutic option in patients with extensive bilateral periventricular nodular heterotopia: a case report. Epileptic Disord. 2020;22:664–668.

Cukiert A. Seizure outcome during bilateral, continuous, thalamic centromedian nuclei deep brain stimulation in patients with generalized epilepsy: a prospective, open-label study. 2020.

Schaper FLW, et al. Deep Brain Stimulation in Epilepsy: A Role for Modulation of the Mammillothalamic Tract in Seizure Control? Neurosurgery. 2020;87:602–610.

Freund BE, et al. Clinical outcome of imaging-based programming for anterior thalamic nucleus deep brain stimulation. J Neurosurg. 2022;1–8. doi:10.3171/2022.7.JNS221116.

Grewal SS, et al. Fast gray matter acquisition T1 inversion recovery MRI to delineate the mammillothalamic tract for preoperative direct targeting of the anterior nucleus of the thalamus for deep brain stimulation in epilepsy. Neurosurg Focus. 2018;45:E6.

Gross RE, et al. Analysis of Deep Brain Stimulation Lead Targeting in the Stimulation of Anterior Nucleus of the Thalamus for Epilepsy Clinical Trial. Neurosurgery. 2021;89:406–412.

Guo W, et al. Defining the optimal target for anterior thalamic deep brain stimulation in patients with drug-refractory epilepsy. J Neurosurg. 2021;134:1054–1063.

Herrera ML, et al. Stimulation of the Anterior Nucleus of the Thalamus for Epilepsy: A Canadian Experience. Can J Neurol Sci. 2021;48:469–478.

Herrman H, et al. Anterior thalamic deep brain stimulation in refractory epilepsy: A randomized, double-blinded study. Acta Neurol Scand. 2018;ane.13047. doi:10.1111/ane.13047.

Kaufmann E, et al. What have we learned from 8 years of deep brain stimulation of the anterior thalamic nucleus? Experiences and insights of a single center. J Neurosurg. 2020;1–10. doi:10.3171/2020.6.JNS20695.

Kerrigan JF, et al. Electrical Stimulation of the Anterior Nucleus of the Thalamus for the Treatment of Intractable Epilepsy. Epilepsia. 2004;45:346–354.

Krishna V, et al. Anterior Nucleus Deep Brain Stimulation for Refractory Epilepsy: Insights Into Patterns of Seizure Control and Efficacious Target. Neurosurgery. 2016;78:802–811.

Lehtimäki K. Outcome based definition of the anterior thalamic deep brain stimulation target in refractory epilepsy. Brain Stimulation. 2016.

Osorio I, et al. High Frequency Thalamic Stimulation for Inoperable Mesial Temporal Epilepsy: HIGH FREQUENCY THALAMIC STIMULATION. Epilepsia. 2007;48:1561–1571.

Passamonti C. Deep brain stimulation in patients with long history of drug resistant epilepsy and poor functional status: Outcomes based on the different targets. Clin Neurol Neurosurg. 2021.

Piacentino M, et al. Anterior thalamic nucleus deep brain stimulation (DBS) for drug-resistant complex partial seizures (CPS) with or without generalization: long-term evaluation and predictive outcome. Acta Neurochir. 2015;157:1525–1532.

Tassigny D, et al. Anterior thalamic nucleus deep brain stimulation for refractory epilepsy: Preliminary results in our first 5 patients. Neurochirurgie. 2020;66:252–257.

Torres Diaz CV, et al. Network Substrates of Centromedian Nucleus Deep Brain Stimulation in Generalized Pharmacoresistant Epilepsy. Neurotherapeutics. 2021;18:1665–1677.

Vakilna YS, et al. Pulvinar neuromodulation for seizure monitoring and network modulation in temporal plus epilepsy. Ann Clin Transl Neurol. 2023;10:1254–1259.

Valent A, et al. Deep brain stimulation of the centromedian thalamic nucleus for the treatment of generalized and frontal epilepsies. 2013.

Velasco AL, et al. Neuromodulation of the Centromedian Thalamic Nuclei in the Treatment of Generalized Seizures and the Improvement of the Quality of Life in Patients with Lennox–Gastaut Syndrome. Epilepsia. 2006;47:1203–1212.

Wang C, et al. Frameless Robot-Assisted Asleep Centromedian Thalamic Nucleus Deep Brain Stimulation Surgery in Patients with Drug-Resistant Epilepsy: Technical Description and Short-Term Clinical Results. Neurol Ther. 2023;12:977–993.

Warren AEL, et al. The Optimal Target and Connectivity for Deep Brain Stimulation in Lennox–Gastaut Syndrome. Ann Neurol. 2022;92.

Yang JC, et al. Centromedian thalamic deep brain stimulation for drug-resistant epilepsy: single-center experience. J Neurosurg. 2022;137:1591–1600.

Costa-Gertrudes R, et al. Anterior Nucleus of Thalamus Deep Brain Stimulation: A Clinical-Based Analysis of the Ideal Target in Drug-Resistant Epilepsy. Stereotact Funct Neurosurg.

Lee KJ, Shon YM, Cho CB. Long-Term Outcome of Anterior Thalamic Nucleus Stimulation for Intractable Epilepsy. Stereotact Funct Neurosurg. 2012;90:379–385.

Khan A, Middlebrooks EH, Freund B, Grewal S, Tatum WO. Is thalamic deep brain stimulation synergistic with vagus nerve stimulation in drug-resistant genetic generalized epilepsy?

Koeppen JA, et al. Electrical Stimulation of the Anterior Thalamus for Epilepsy: Clinical Outcome and Analysis of Efficient Target. 2018.

Miron G, Strauss I, Fried I, Fahoum F. Anterior thalamic deep brain stimulation in epilepsy patients refractory to vagus nerve stimulation: A single center observational study. Epilepsy Behav Rep. 2022;20:100563.

Poulen G, Rolland A, Chan-Seng E, Sanrey E. Microendoscopic transventricular deep brain stimulation of the anterior nucleus of the thalamus as a safe treatment in intractable epilepsy: A feasibility study. Rev Neurol. 2022.

Scherer M. Desynchronization of temporal lobe theta-band activity during effective anterior thalamus deep brain stimulation in epilepsy. 2020.

Sitnikov A, Grigoryan Y, Mishnyakova L. Bilateral stereotactic lesions and chronic stimulation of the anterior thalamic nuclei for treatment of pharmacoresistant epilepsy. Surg Neurol Int. 2018;9:137.

Sobstyl M, et al. Clinical efficacy and safety of anterior thalamic deep brain stimulation for intractable drug-resistant epilepsy. Epilepsy Res. 2023;195:107199.

Tong X. Analysis of power spectrum and phase lag index changes following deep brain stimulation of the anterior nucleus of the thalamus in patients with drug-resistant epilepsy: A retrospective study. 2022.

Wang YC. Probing circuit of Papez with stimulation of anterior nucleus of the thalamus and hippocampal evoked potentials. Epilepsy Res. 2020.

Chua MMJ, et al. Initial case series of a novel sensing deep brain stimulation device in drug-resistant epilepsy and consistent identification of alpha/beta oscillatory activity: A feasibility study.

Bondallaz P. Electrode location and clinical outcome in hippocampal electrical stimulation for mesial temporal lobe epilepsy. 2013.

Boon P, et al. Deep Brain Stimulation in Patients with Refractory Temporal Lobe Epilepsy. 2007;48.

Cukiert A, Cukiert CM, Burattini JA, Mariani PP. Long-term seizure outcome during continuous bipolar hippocampal deep brain stimulation in patients with temporal lobe epilepsy with or without mesial temporal sclerosis: An observational, open-label study.

Jin H. Hippocampal deep brain stimulation in nonlesional refractory mesial temporal lobe epilepsy. 2016.

Saucedo-Alvarado PE, et al. Optimizing deep brain stimulation for the treatment of drug-resistant temporal lobe epilepsy: a pilot study.

Tellez-Zenteno JF, McLachlan RS, Parrent A, Kubu CS, Wiebe S. Hippocampal electrical stimulation CME in mesial temporal lobe epilepsy.

Velasco AL, et al. Electrical Stimulation of the Hippocampal Epileptic Foci for Seizure Control: A Double-Blind, Long-Term Follow-Up Study. 2007;48.

Son BC, et al. Clinical Outcome of Patients with Deep Brain Stimulation of the Centromedian Thalamic Nucleus for Refractory Epilepsy and Location of the Active Contacts. Stereotact Funct Neurosurg. 2016;94:187–197.

Vilela-Filho O, Arruda FM. A new strategy for treating drug-resistant focal aware seizures: thalamic specific nuclei deep brain stimulation. Illustrative case.

Benedetti-Isaac JC, et al. Seizure frequency reduction after posteromedial hypothalamus deep brain stimulation in drug-resistant epilepsy associated with intractable aggressive behavior. 2015.

Cui Z, et al. Long-term efficacy of deep brain stimulation of the subthalamic nucleus in patients with pharmacologically intractable epilepsy: A case series of six patients.

Sobstyl M. Deep brain stimulation in a patient with progressive myoclonic epilepsy and ataxia due to potassium channel mutation (MEAK). A case report and review of the literature. 2023.

Wang X, et al. Long-term outcome of unilateral deep brain stimulation of the subthalamic nucleus for a patient with drug-resistant focal myoclonic seizure. Ann Transl Med. 2020;8.

Deep Brain Stimulation of Two Unconventional Targets in Refractory Non-Resectable Epilepsy. Stereotact Funct Neurosurg.

Van Gompel JJ, et al. Anterior nuclear deep brain stimulation guided by concordant hippocampal recording. FOC. 2015;38:E9.

Vilela-Filho O, Ragazzo PC, Goulart LC, Arruda F, Arruda ML. Ventral intermediate nucleus deep brain stimulation for treatment-resistant focal aware motor seizures: illustrative case.

Wille C, et al. Chronic high-frequency deep-brain stimulation in progressive myoclonic epilepsy in adulthood—Report of five cases: DBS in Progressive Myoclonic Epilepsy. Epilepsia. 2011;52:489–496.

Yan H, et al. Deep brain stimulation for patients with refractory epilepsy: nuclei selection and surgical outcome. Front Neurol. 2023;14:1169105.

Zheng H. Deep Brain Stimulation of Anterior Thalamic Nucleus for Treatment of Patient with Tuberous Sclerosis-Related Refractory Epilepsy. World Neurosurg. 2020.

Schmitt FC, et al. Safety and feasibility of nucleus accumbens stimulation in five patients with epilepsy. J Neurol. 2014.

Imbach LL. Anticonvulsive effect of anterior thalamic deep brain stimulation in super-refractory status epilepticus crucially depends on active stimulation zone—A single case observation. 2019.

Järvenpää S, Lehtimäki K, Rainesalo S, Möttönen T, Peltola J. Improving the effectiveness of ANT DBS therapy for epilepsy with optimal current targeting.

Kim SH. Long-term follow-up of anterior thalamic deep brain stimulation in epilepsy: An 11-year, single center experience. 2017.

Kim HY, et al. Modification of electrophysiological activity pattern after anterior thalamic deep brain stimulation for intractable epilepsy: report of 3 cases.

Lee CY. Successful Treatment of Refractory Status Epilepticus Using Anterior Thalamic Nuclei Deep Brain Stimulation. World Neurosurg. 2017.

Lim SN, et al. Low and High Frequency Hippocampal Stimulation for Drug-Resistant Mesial Temporal Lobe Epilepsy. 2016.

Hupalo M. Intracerebral electroencephalography in targeting anterior thalamic nucleus for deep brain stimulation in refractory epilepsy. Neurol Neurochir Pol. 2018.

Marras CE, Messina G, Franzini A. Deep brain stimulation for the treatment of drug-refractory epilepsy in a patient with a hypothalamic hamartoma. Neurosurg Focus. 2011;30.

Khan S, Carter M, Gill SS. High frequency stimulation of the mamillothalamic tract for the treatment of resistant seizures associated with hypothalamic hamartoma. 2009.

Martin RA, Cukiert A, Blumenfeld H. Short-term changes in cortical physiological arousal measured by electroencephalography during thalamic centromedian deep brain stimulation.

Middlebrooks EH, et al. Differences in functional connectivity profiles as a predictor of response to anterior thalamic nucleus deep brain stimulation for epilepsy: a hypothesis for the mechanism of action and a potential biomarker for outcomes. Neurosurg Focus. 2018;45:E7.

Middlebrooks EH, Jain A, Okromelidze L, Lin C, Westerhold EM. Acute Brain Activation Patterns of High- Versus Low-Frequency Stimulation of the Anterior Nucleus of the Thalamus During Deep Brain Stimulation for Epilepsy.

Peltola J. Stimulation Induced Electrographic Seizures in Deep Brain Stimulation of the Anterior Nucleus of the Thalamus Do Not Preclude a Subsequent Favorable Treatment Response. Front Neurol. 2018;9.

Olaciregui Dague K, Witt JA, Von Wrede R, Helmstaedter C, Surges R. DBS of the ANT for refractory epilepsy: A single center experience of seizure reduction, side effects and neuropsychological outcomes. Front Neurol. 2023;14:1106511.

Peltola J, et al. Deep Brain Stimulation of the Anterior Nucleus of the Thalamus in Drug-Resistant Epilepsy in the MORE Multicenter Patient Registry. Neurology. 2023;100.

Picillo M, Rohani M, Lozano AM, Fasano A. Two indications, one target: Concomitant epilepsy and Tourettism treated with Centromedian/parafascicular thalamic stimulation. Brain Stimul. 2017;10:711–713.

Sandoval-Bonilla BA, Codeiro JG, Cosio JF. Adequate control of seizures in a case of lead migration and neuromodulation of the posterior Sylvian junction: A case report.

Sarica C. Blood oxygen level-dependent (BOLD) response patterns with thalamic deep brain stimulation in patients with medically refractory epilepsy. 2021.

Shan M. Deep brain stimulation of the subthalamic nucleus for a patient with drug resistant juvenile myoclonic epilepsy: 1 year follow-up. Neurol Sci.

Shon YM, et al. Effect of Chronic Deep Brain Stimulation of the Subthalamic Nucleus for Frontal Lobe Epilepsy: Subtraction SPECT Analysis. Stereotact Funct Neurosurg.

Suresh S, et al. Case report: Nocturnal low-frequency stimulation of the centromedian thalamic nucleus improves sleep quality and seizure control. Front Hum Neurosci.

Wu C, et al. Variations in Thalamic Anatomy Affect Targeting in Deep Brain Stimulation for Epilepsy. Stereotact Funct Neurosurg. 2016;94:387–396.

Yang AI, Isbaine F, Alwaki A, Gross RE. Multitarget deep brain stimulation for epilepsy.

Gouveia FV. Case report: 5 Years follow-up on posterior hypothalamus deep brain stimulation for intractable aggressive behaviour associated with drug-resistant epilepsy. Brain Stimul. 2021.

Harrison DJ, Oushy S, Gregg NM, Lundstrom BN, Van Gompel JJ. Stereotactic depth electrode placement for chronic subthreshold cortical stimulation: surgical technique video. 2024;11.

Aiello G. Functional network dynamics between the anterior thalamus and the cortex in deep brain stimulation for epilepsy.

Sweeney-Reed CM. Thalamic interictal epileptiform discharges in deep brain stimulated epilepsy patients. J Neurol. 2016.

Cukiert A, Cukiert CM, Burattini JA, Lima AM. Seizure Outcome After Battery Depletion in Epileptic Patients Submitted to Deep Brain Stimulation. 2015.

Torres CV, Pastor J, García-Navarrete E, García-Camba E. Long-term results of posteromedial hypothalamic deep brain stimulation for patients with resistant aggressiveness. J Neurosurg. 2013.

Kowski AB, Holtkamp M, Schmitt FC. Nucleus accumbens stimulation in partial epilepsy—A randomized controlled case series. 2015.

Sobstyl M. Deep brain stimulation of the subiculum in the treatment for refractory temporal lobe epilepsy due to unilateral mesial temporal lobe sclerosis. 2024.

Jordán Z. Epileptiform discharges in the anterior thalamus of epilepsy patients. Open Access.

Satzer D, et al. Ambulatory Local Field Potential Recordings from the Thalamus in Epilepsy: A Feasibility Study. 2024.

Yang JC, et al. Anterior nucleus of the thalamus deep brain stimulation vs temporal lobe responsive neurostimulation for temporal lobe epilepsy.

Koubeissi MZ. Low-frequency stimulation of a fiber tract in bilateral temporal lobe epilepsy. 2022.

Piacentino M. Hippocampal deep brain stimulation: persistent seizure control after bilateral extra-cranial electrode fracture. Neurol Sci. 2018.

Hartl E. Seizure reductions outlast DBS explantation. Brain Stimul. 2018.

Lim SN, et al. Electrical Stimulation of the Anterior Nucleus of the Thalamus for Intractable Epilepsy: A Long-term Follow-up Study. 2007;48.

Handforth A, DeSalles AAF, Krahl SE. Deep Brain Stimulation of the Subthalamic Nucleus as Adjunct Treatment for Refractory Epilepsy.

Cukiert A. Combined Neuromodulation (Vagus Nerve Stimulation and Deep Brain Stimulation) in Patients With Refractory Generalized Epilepsy: An Observational Study. 2022.

Boongird A, et al. Deep Brain Stimulation of Anterior Thalamic Nuclei for Intractable Epilepsy in Thailand: Case Report. 2016;99.

Velasco F, et al. Electrical Stimulation of the Centromedian Thalamic Nucleus in Control of Seizures: Long-Term Studies. Epilepsia. 1995;36:63–71.

Fisher RS, et al. Placebo-Controlled Pilot Study of Centromedian Thalamic Stimulation in Treatment of Intractable Seizures. Epilepsia. 1992;33:841–851.

Velasco F, et al. Double-Blind, Randomized Controlled Pilot Study of Bilateral Cerebellar Stimulation for Treatment of Intractable Motor Seizures. Epilepsia. 2005;46:1071–1081.

Capecci M. Chronic bilateral subthalamic stimulation after anterior callosotomy in drug-resistant epilepsy: Long-term clinical and functional outcome of two cases.

Stavropoulos I. Low frequency centromedian thalamic nuclei deep brain stimulation for the treatment of super refractory status epilepticus: A case report and a review of the literature. Brain Stimul. 2021.

Hect JL, Fernandez LD, Welch WP, Abel TJ. Deep brain stimulation of the centromedian thalamic nucleus for the treatment of FIRES. Epilepsia Open. 2022;7:187–193.

Valentín A. Centromedian thalamic nuclei deep brain stimulation in refractory status epilepticus.

Lehtimäki K, et al. Successful management of super-refractory status epilepticus with thalamic deep brain stimulation. Ann Neurol. 2017;81:142–146.

Cukiert A, et al. Centro-median stimulation yields additional seizure frequency and attention improvement in patients previously submitted to callosotomy. Seizure. 2009;18:588–592.

Sa M, et al. Centromedian thalamic nuclei deep brain stimulation and Anakinra treatment for FIRES - Two different outcomes. Eur J Paediatr Neurol. 2019;23:749–754.

Andrade DM, et al. Long-term follow-up of patients with thalamic deep brain stimulation for epilepsy. Neurology. 2006;66:1571–1573.

Gillinder L, et al. Refractory epilepsy secondary to anti-GAD encephalitis treated with DBS post SEEG evaluation: a novel case report based on stimulation findings. Epileptic Disord. 2018;20:451–456.

di Giacopo A, et al. Selective deep brain stimulation in the substantia nigra reduces myoclonus in progressive myoclonic epilepsy. Epileptic Disord. 2019;21:283–288.

Chabardès S, et al. Deep brain stimulation in epilepsy with particular reference to the subthalamic nucleus. Epileptic Disord. 2002;4 Suppl 3:S83-93.

McLachlan RS, Pigott S, Tellez-Zenteno JF, Wiebe S, Parrent A. Bilateral hippocampal stimulation for intractable temporal lobe epilepsy: impact on seizures and memory. Epilepsia. 2010;51:304–307.

Yuan L, et al. Deep brain stimulation of the anterior nucleus of the thalamus in a patient with super-refractory convulsive status epilepticus. Epileptic Disord. 2019;21:379–384.

Wang S. Long-term efficacy and cognitive effects of bilateral hippocampal deep brain stimulation in patients with drug-resistant temporal lobe epilepsy. Neurol Sci. 2021.

Vázquez-Barrón D, Cuéllar-Herrera M, Velasco F, Velasco AL. Electrical Stimulation of Subiculum for the Treatment of Refractory Mesial Temporal Lobe Epilepsy with Hippocampal Sclerosis: A 2-Year Follow-Up Study. Stereotact Funct Neurosurg. 2021;99:40–47.

104.	Vilela-Filho, O., Ragazzo, P. C., Goulart, L. C., Arruda, F. & Arruda, M. L. Ventral intermediate nucleus deep brain stimulation for treatment-resistant focal aware motor seizures: illustrative case.

105.	Deep Brain Stimulation of Two Unconventional Targets in Refractory Non-Resectable Epilepsy. *Stereotact Funct Neurosurg*.

106.	Yang, A. I., Isbaine, F., Alwaki, A. & Gross, R. E. Multitarget deep brain stimulation for epilepsy. *J Neurosurg* **140**, 210–217 (2024).

107.	Schmitt, F. C. *et al.* Safety and feasibility of nucleus accumbens stimulation in ﬁve patients with epilepsy. *J Neurol* (2014).

108.	Khan, S., Carter, M. & Gill, S. S. High frequency stimulation of the mamillothalamic tract for the treatment of resistant seizures associated with hypothalamic hamartoma. (2009).

109.	Benedetti-Isaac, J. C. *et al.* Seizure frequency reduction after posteromedial hypothalamus deep brain stimulation in drug‐resistant epilepsy associated with intractable aggressive behavior. (2015).

110.	Gouveia, F. V. Case report: 5 Years follow-up on posterior hypothalamus deep brain stimulation for intractable aggressive behaviour associated with drug-resistant epilepsy. *Brain Stimulation* (2021).

111.	Torres, C. V., Pastor, J., García-Navarrete, E. & García-Camba, E. Long-term results of posteromedial hypothalamic deep brain stimulation for patients with resistant aggressiveness. *J Neurosurg* **119**, (2013).

112.	Vilela-Filho, O. & Arruda, F. M. A new strategy for treating drug-resistant focal aware seizures: thalamic specific nuclei deep brain stimulation. Illustrative case.

113.	Yan, H. *et al.* Deep brain stimulation for patients with refractory epilepsy: nuclei selection and surgical outcome. *Front. Neurol.* **14**, 1169105 (2023).

1. Percentages and averages are reported as calculated based on available data. Missing data was not imputed.
